# Impact of active case finding for tuberculosis with mass chest X-ray screening in Glasgow, Scotland, 1950-1963: an epidemiological analysis of historical data

**DOI:** 10.1101/2024.07.25.24310967

**Authors:** Peter MacPherson, Helen R Stagg, Alvaro Schwalb, Hazel Henderson, Alice E Taylor, Rachael M Burke, Hannah M Rickman, Cecily Miller, Rein MGJ Houben, Peter J Dodd, Elizabeth L Corbett

## Abstract

**Background:** Community active case finding for tuberculosis was widely implemented in Europe and North America between 1940-1970, when incidence was comparable to many present-day high-burden countries. Using an interrupted time series analysis, we analysed the effect of the 1957 Glasgow mass chest X-ray campaign to inform contemporary approaches to screening.

**Methods and findings:** Case notifications for 1950-1963 were extracted from public health records and linked to demographic data. We fitted Bayesian multilevel regression models to estimate annual relative case notification rates (CNR) during and after a mass screening intervention implemented over five weeks in 1957 compared to the counterfactual scenario where the intervention had not occurred. We additionally estimated case detection ratios and incidence. From 11/3/1957-12/4/1957, 714,915 people (622,349 of 819,301 [76.0%] resident adults ≥15 years) were screened with miniature chest X-ray; 2,369 (0.4%) were diagnosed with tuberculosis. Pre-intervention (1950-1956), pulmonary CNRs were declining at 2.3% per year from a CNR of 222/100,000 in 1950. With the intervention in 1957, there was a doubling in the pulmonary CNR (RR: 1.95, 95% uncertainty interval [UI]: 1.80-2.11), and 36% decline in the year after (RR: 0.64, 95%UI: 0.59-0.71). Post-intervention (1958-1963) annual rates of decline (5.4% per year) were greater (RR: 0.77, 95%UI: 0.69-0.85), and there were an estimated 4,656 (95%UI: 3,670-5,725) pulmonary case notifications averted due to the intervention. Effects were consistent across all city wards and notifications declined substantially in young children (0-5 years) with the intervention.

**Conclusions:** A single, rapid round of mass screening with chest X-ray (probably the largest ever conducted) resulted in a major and sustained reduction in tuberculosis case notifications. Contemporary high-burden settings may achieve similar benefits with high-intensity screening.

## Introduction

Tuberculosis has been a scourge of humankind for millennia^1^. From the late 18^th^ to the early 20^th^centuries, improvements in housing, nutrition, and air quality^2^ – as well as progressive improvements in tuberculosis care, treatment, and prevention^3^ – yielded substantial reductions in incidence and mortality in countries such as the United Kingdom^4^.

By the 1940s, it was recognised that additional measures would be required to stem the source of *Mycobacterium tuberculosis* infections and prevent tuberculosis disease^5^. In line with other European and American cities in the 1940-60s^6^, Scotland implemented active case finding (ACF) in communities through mass chest X-ray screening campaigns^7^. To our knowledge, the largest single site tuberculosis ACF intervention ever implemented globally was in Glasgow, Scotland, in 1957^6^. By the end of the Second World War, Glasgow had extremely high levels of social deprivation and nearly one out of every thousand people resident in Glasgow died of pulmonary tuberculosis annually^8^. Whereas incidence of tuberculosis and mortality had declined substantially in other major UK cities, progress in Glasgow had lagged considerably behind^9^.

In 1974 the World Health Organization (WHO) recommended *“the policy of indiscriminate tuberculosis case finding by mobile mass radiography should now be abandoned”* due to the declining diagnostic yield from screening programmes^10^. Yet currently many countries in Africa and Asia have tuberculosis epidemiological indicators similar to those of Europe and North America in the 1940s and 1950s^11^. This suggests that the experiences of ACF in cities like Glasgow are highly informative for the modern day. Indeed, the WHO made a conditional recommendation in favour of ACF for communities where the prevalence of undiagnosed pulmonary tuberculosis was >1,000 per 100,000 in 2013^12^, lowering this threshold to a prevalence of >500 per 100,000 in 2021^13^.

Action is needed to implement high-impact interventions that can change the trajectory of tuberculosis epidemics in high-burden nations. Despite renewal of interest in ACF^14^ – and more recent trial evidence^15,16^ – there remains considerable uncertainty over how and where ACF should be implemented, and what effects can be expected to be achieved^17^. Using comprehensive historical records, we therefore set out to estimate the impact of the 1957 Glasgow mass tuberculosis screening intervention to inform contemporary approaches to ACF in high tuberculosis burden countries.

## Methods

### Mass miniature X-ray screening intervention

The Glasgow mass X-ray campaign was coordinated by the Corporation of Glasgow, the Western Regional Hospital Board, the Scottish Information Office, and the Department of Health for Scotland, and X-ray screening took place between the 11^th^ of March and 12^th^ April 1957^18^. A total of 37 mobile miniature X-ray units with radiographers were seconded from cities across the UK, and the campaign was supported by ∼12,000 Glasgow volunteers. Across the city, screening activities were divided into six sections that aligned with five geographical city divisions plus a further section focused on activities within the city centre and business area.

Prior to the campaign substantial publicity and public engagement was undertaken, including: house-to-house visits within each city ward to distribute X-ray invitation cards; press and cinema advertising; poster displays; church services; distribution of leaflets, banners and stickers; loudspeaker vans and an illuminated tramcart; pavement stencils; aeroplane banner advertising; two specially-commissioned campaign songs that were played on the radio and at football matches; and information printed on municipal letterheads.

All people who underwent chest X-ray received a badge, and randomly selected people wearing badges within Divisions received small gifts such as chocolates, chickens, and cigarettes. During each week of the campaign, larger gifts (refrigerators, televisions, washing machines, holidays, furniture, a car) were distributed by selecting X-ray cards at random, and the 100,000^th^200,000^th^ and 250,000^th^ person X-rayed received additional gifts.

Mobile X-ray units were situated in department stores, churches, schools, and other municipal buildings. People aged 15 years or older were invited to receive chest X-ray, regardless of the presence or absence of symptoms, with miniature films interpreted by medical officers. Where there was “significant radiological abnormality requiring further investigation or supervision”, people were recalled for a large film at a central X-ray site, and assessment/further investigation by physicians at five city hospitals chest clinics (one per Division, including microbiological testing of sputum) and tuberculosis treatment (if required).

### Setting and Population

For this analysis, we used the municipal boundaries of Glasgow City as defined in 1951. Wards are electoral boundaries used in UK national parliamentary elections, and Glasgow City was divided into 37 wards within five divisions (North, East, Central, South-East, South-West). To obtain spatial boundaries, we digitised a scale 1951 Post Office Directory map held in the City of Glasgow Archives using QGIS software (Supplemental Figure S1).

We extracted overall city and ward-specific annual population estimates as reported in Medical Officer of Health reports for Glasgow between 1950 and 1963^19^. In reports, population estimates were based on national censuses (1951 and 1961), updated annually by the Medical Officer of Health based on linear projections of ratios of census populations to registered voters. For this analysis, we used two population denominators available: a) *total population* (all people identified as usually resident within the City of Glasgow, including people in long-term institutional care and sailors stationed on ships of the Royal Navy at sea or in ports abroad); and b) the *population excluding people in institutional care or shipping.* In reports, population denominators were adjusted for each year to account for city-wide and ward-specific trends. Population estimates stratified by age and sex were not available for each ward.

### Tuberculosis notifications

For each year between 1950 and 1963, we extracted from annual Medical Officer of Health Reports for the City of Glasgow^19^ overall numbers of tuberculosis notifications (stratified by pulmonary and extra-pulmonary status, and separately by age group and sex), and the ward-specific notifications (stratified by pulmonary and extra-pulmonary status, and sex). In 1962 and 1963, extra-pulmonary cases were not reported by ward due to small numbers.

### Statistical methods

We calculated the annual pulmonary and extra-pulmonary tuberculosis case notification rate for the City of Glasgow using overall numbers of notifications and the *total population* denominator and scaled these data per 100,000 people. We additionally calculated the ward-specific pulmonary and extrapulmonary case notification rates using the *population excluding people in institutional care or shipping* as the denominator, as cases occurring among institutionalised people or in shipping were not allocated to wards.

To investigate the impact of the 1957 active case finding intervention, we constructed multi-level Bayesian interrupted time series regression models (Supplemental Equation 1) to estimate annual pulmonary tuberculosis case notification rates and extra-pulmonary tuberculosis notification rates. Models included terms for a “level-change” in 1957 to capture the immediate effect of the intervention and a “slope change” to estimate changing rates before and after the intervention. To account for over-dispersion, we used the negative binomial distributional family; priors were weakly informative and assessed by inspecting plots of joint prior distributions. Models were fit using the ‘brms’ interface to Stan in R^20^, and model convergence was assessed using *Ȓ* statistics, effective sample size measures, trace plots of chains, and posterior predictive plots comparing observed data to simulated data from the empirical cumulative distribution function.

We drew 4000 samples from model posteriors and summarised these (using means and quantile-based 95% uncertainty intervals [UI]). To investigate the impact of the ACF intervention overall and by ward, we predicted counterfactual case notification rates for 1958-1963 based on a linear projection of trends from the pre-ACF period (1950-1956) under the scenario where the ACF intervention had not happened. We then estimated: i) the relative case notification rate in 1957 compared to the counterfactual for the same year (*“peak effect”*); ii) the *“level effect”* (relative rate in 1958 vs. counterfactual for 1958); and iii) the *“slope effect”* (relative annual change in case notifications in 1958 to 1963 compared to the counterfactual scenario). Pairwise correlations between posterior draws for the peak effect, step effect, and slope effect were plotted overall, and by ward. Using model predictions, we calculated the number of tuberculosis case notifications that were averted in the post-ACF period (1958-1963) compared to that predicted by the counterfactual scenario. We additionally investigated whether the impact of the ACF intervention differed by age and sex by fitting a model to estimate the annual counts of pulmonary tuberculosis case notifications using a Poisson distribution, and summarised as above. All analysis was conducted using R version 4.2.3 (R Core Team, Vienna).

### Estimates of case detection and incidence

We estimated the incidence and case detection ratio (CDR; percentage of estimated new pulmonary TB cases notified annually) for each ward using an equilibrium competing hazards model and the assumption that the excess notified cases (*N_during_ – N_pre_)* were prevalent during the intervention, and detected with a coverage (*cov)* taken to be 76% based on screening uptake (Supplemental Methods, Page 4).

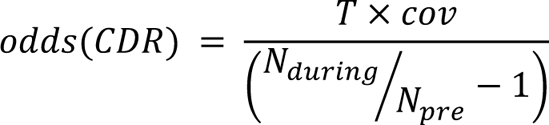

Here, *T* is the average duration of tuberculosis disease in the absence of detection and treatment, taken to be 3 years^21^. We examined the empirical correlations across wards between CDR, pre-ACF incidence, and ACF impact quantified as relative decrease in notifications pre/post-ACF.

### Ethical statement and data availability

Ethical approval was not required for this analysis. Data and code to reproduce analysis are available at https://github.com/petermacp/glasgow-cxr.

## Results

### Population

The total population of Glasgow City was 1.1 million in 1950, declining to just over 1 million by 1963 (Table 1). Ward populations ranged from 16,321 people (Knightswood) to 44,595 (Dalmarnock) in 1950. There was considerable variation in the change in population by 1963, with some wards having substantial population increases (Provan: +312%), and others declining (Exchange: -57%) (Supplemental Figure S2). Between 1950 and 1963, we included a total of 14,649,693 person-years of follow-up (*population excluding people in institutional care or shipping)* in regression models.

**Table 1:** Population, and tuberculosis notifications in Glasgow City, 1950-1963.

| Year | Total estimated population* | Pulmonary tuberculosis notifications | Pulmonary tuberculosis case notification rate (per 100,000) | Extra-pulmonary tuberculosis notifications | Extra-pulmonary tuberculosis notification rate (per 100,000) | Total tuberculosis notifications | Total tuberculosis case notification rate (per 100,000) | Percent of tuberculosis notifications that were pulmonary |
| --- | --- | --- | --- | --- | --- | --- | --- | --- |
| 1950 | 1,100,000 | 2,446 | 222.4 | 369 | 33.5 | 2,815 | 255.9 | 86.9% |
| 1951 | 1,089,767 | 2,207 | 202.5 | 355 | 32.6 | 2,562 | 235.1 | 86.1% |
| 1952 | 1,086,800 | 2,264 | 208.3 | 301 | 27.7 | 2,565 | 236.0 | 88.3% |
| 1953 | 1,085,000 | 2,368 | 218.2 | 295 | 27.2 | 2,663 | 245.4 | 88.9% |
| 1954 | 1,084,700 | 2,201 | 202.9 | 241 | 22.2 | 2,442 | 225.1 | 90.1% |
| 1955 | 1,085,100 | 2,181 | 201.0 | 278 | 25.6 | 2,459 | 226.6 | 88.7% |
| 1956 | 1,083,500 | 2,024 | 186.8 | 193 | 17.8 | 2,217 | 204.6 | 91.3% |
| 1957<sup>†</sup> | 1,079,800 | 3,925 | 363.5 | 172 | 15.9 | 4,097 | 379.4 | 95.8% |
| 1958 | 1,078,400 | 1,340 | 124.3 | 167 | 15.5 | 1,507 | 139.7 | 88.9% |
| 1959 | 1,075,800 | 1,159 | 107.7 | 120 | 11.2 | 1,279 | 118.9 | 90.6% |
| 1960 | 1,064,700 | 1,092 | 102.6 | 109 | 10.2 | 1,201 | 112.8 | 90.9% |
| 1961 | 1,053,100 | 1,021 | 97.0 | 137 | 13.0 | 1,158 | 110.0 | 88.2% |
| 1962 | 1,044,500 | 927 | 88.8 | 117 | 11.2 | 1,044 | 100.0 | 88.8% |
| 1963 | 1,029,147 | 863 | 83.9 | 116 | 11.3 | 979 | 95.1 | 88.2% |

\*Total populations and case notifications, including those in institutional care and shipping
<sup>†</sup>Mass miniature X-ray campaign in 1957

Across each year in Glasgow City, there was a greater number of females than males, with the sex ratio slightly declining from 1:1.10 in 1950 to 1.08 in 1963. The difference in population sex ratio was driven by fewer men compared to women aged 45 years or older (Supplemental Figure S3).

### The Glasgow mass miniature chest X-ray campaign

Between 11^th^ March and 12^th^ April 1957, a total of 714,915 people underwent miniature chest X-ray in the campaign. Of these, 19,466 (2.7%) were <15 years, and 73,100 (10.2%) were not resident in Glasgow, leaving 622,349 adult Glasgow-resident participants, 76.0% of the estimated 819,301 adult resident population. A greater percentage of female adult residents (340,474/437,588, 77.8%) than male (281,875/381,713, 73.8%) underwent chest X-ray, and uptake was highest in younger age groups (Supplemental Figure S4). A total of 30,506 people (4.3% of all X-rayed) were recalled for full film, with 652, (2.1%) not attending; 13,900 (45.6%) were subsequently assessed at chest clinics. In total, 2,565 participants were diagnosed as having new active tuberculosis requiring treatment, with 33 of these <15 years and 196 non-resident, resulting in 2,369/622,349 (0.4%) Glasgow resident cases detected; approximately 65% were treated as outpatients. A further 1,556 people of all ages were notified with tuberculosis through routine systems during 1957, meaning that the ACF campaign accounted for 60% of all notifications in that year. Of the 2,369 adult Glasgow residents diagnosed with tuberculosis due to screening, 58.5% (1,387) were male, and prevalence was highest in older age groups; 523 (22%) were bacteriologically-confirmed by isolation of *M. tuberculosis*. In the year of the campaign, tuberculosis notifications were lower in young children (0-5 years) compared to the years before, and a greater percentage of TB cases were in older age groups; this was maintained in the post-ACF period (Supplemental Figure S5). A substantial burden of other disease was identified through the miniature chest X-ray campaign, including lung cancer (n=327), pulmonary fibrosis (n=1,279), and cardiac abnormalities (n=1,072).

### Tuberculosis case notifications prior to active case finding intervention

In 1950, a total of 2,815 people were notified with tuberculosis in Glasgow City, giving a case notification rate of 255.9 per 100,000. Of these 87% were pulmonary notifications (n=2,446, case notification rate: 222.4 per 100,000), and 13% were extra-pulmonary (n=369, case notification rate: 33.5 per 100,000). Pulmonary tuberculosis case notifications rates varied by ward, with the highest case notification rate in 1950 in Provan ward (68/19,297, 352.4 per 100,000), and the lowest in Camphill ward (16/23,630, 67.8 per 100,000) – Figure 1.

**Figure 1:**
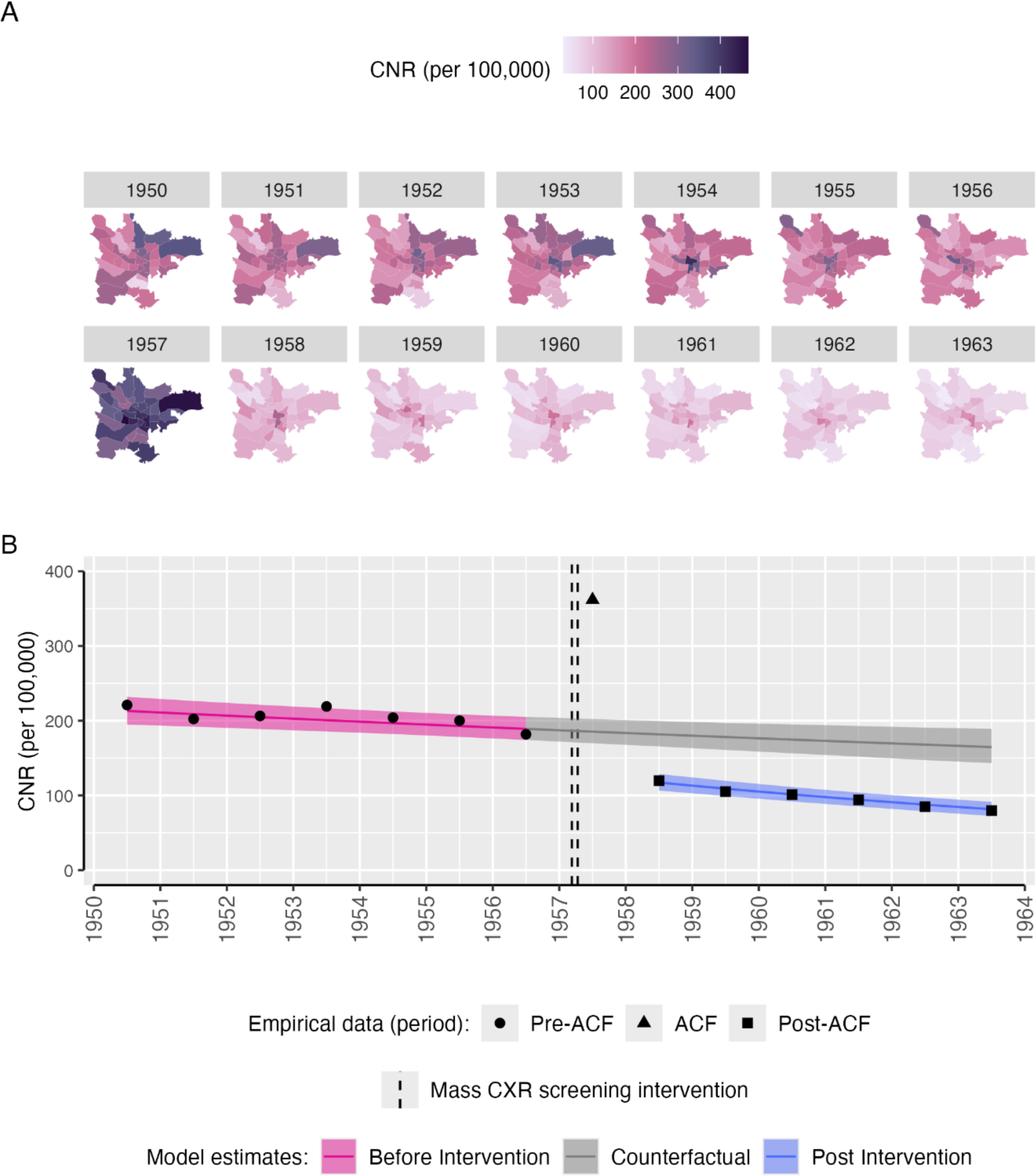
Pulmonary tuberculosis case notification rates in Glasgow City, 1950-1963. **A** Pulmonary tuberculosis case notification rates (per 100,000 population), by Glasgow City ward. ward names can be seen in Supplemental Figure S1. **B** Empirical (points) and modelled (pink and blue bands) pulmonary tuberculosis case notification rates (per 100,000 population), with counterfactual of no ACF intervention (grey bands). The mass miniature X-ray active case finding campaign occurred between dashed lines (11th March-12th April 1957). Points are empirical data based on total population estimates and numbers of pulmonary tuberculosis notifications reported to the Glasgow Medical Officer of Health in 1950-1963. Coloured bands are 95% uncertainty intervals, estimated from a Bayesian negative binomial interrupted time-series regression model. ACF: active case finding. CNR: case notification rate.

Between 1950 and 1956 (pre-ACF period), there was a slow linear decline in pulmonary tuberculosis case notification rates overall (equivalent to a 2.3% per year reduction), and in most wards (Figure 1). Wards in the Central and East Districts tended to have higher notification rates than other parts of the city, with some wards showing a flat, or increasing, trend (Supplemental Figure S6).

### Impact of active case finding campaign on pulmonary tuberculosis notifications

Inspection of trace plots and model diagnostics showed regression models converged well (Supplemental Figure S7 and Table S1).

In 1957, when the mass miniature X-ray active case finding intervention was undertaken, there was a doubling in the case notification rate for Glasgow City as a whole, increasing from 186.8 per 100,000 in 1956 to 363.5 per 100,000 in 1957. In the interrupted time series regression model, comparing 1957 (ACF intervention year) to the counterfactual scenario there was a 1.95-times (95% UI: 1.80 to 2.11) increase in the pulmonary tuberculosis case notification rate across the city (*‘peak effect’)*. At the ward level, there were substantial increases in the pulmonary tuberculosis case notification rate in all wards, with relative rates ranging from 1.73 (95% UI: 1.38 to 2.05) in Gorbals to 2.23 (95% UI: 1.81 to 2.85) in Camphill (Figure 2).

**Figure 2:**
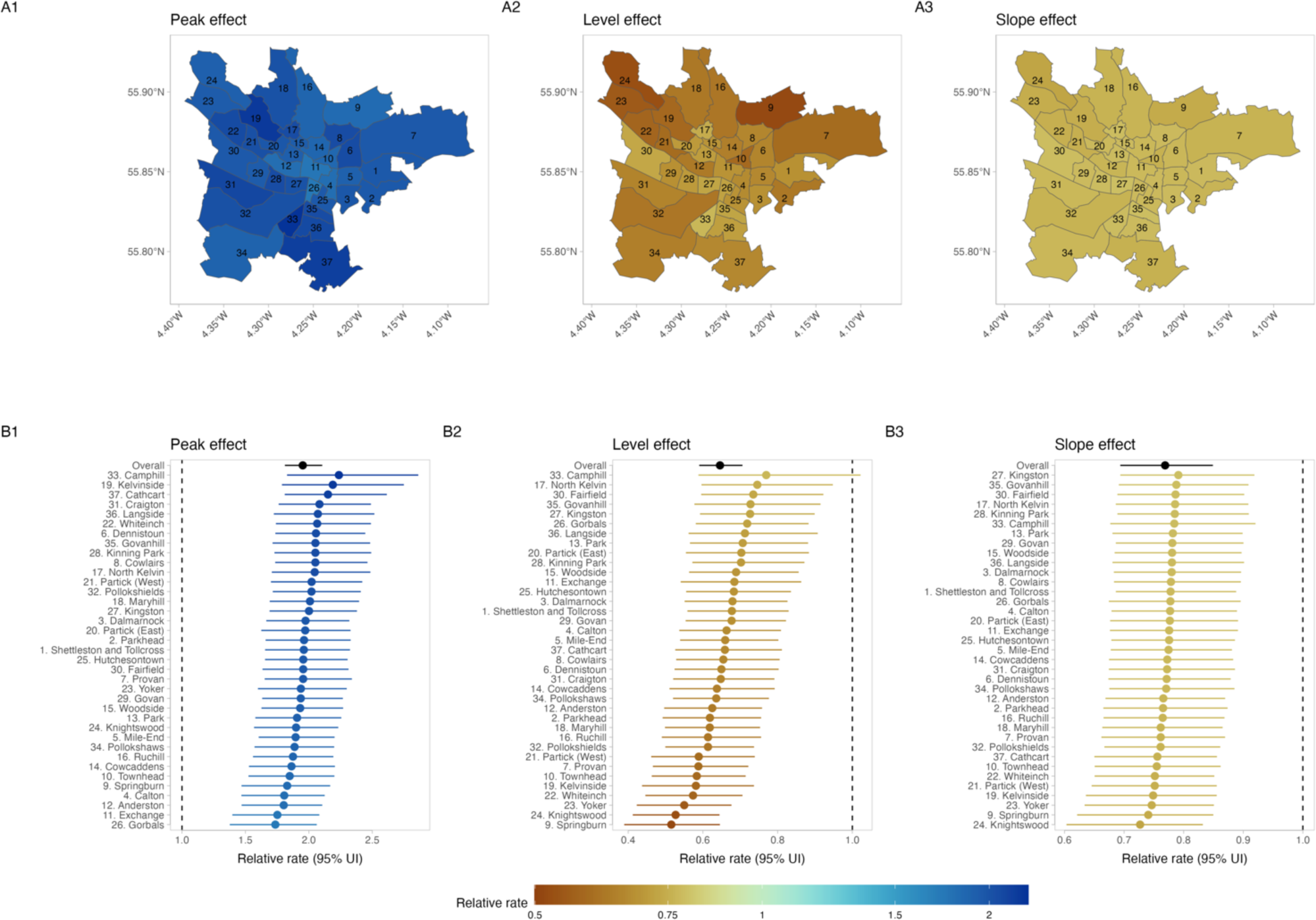
Impact of mass chest X-ray screening on pulmonary tuberculosis case notification rates, overall, and by ward A1. Mean posterior relative pulmonary tuberculosis case notification rate in 1957 vs. counterfactual (“peak effect”). **A2** Mean posterior relative pulmonary tuberculosis case notification rate in 1958 vs. counterfactual (“level effect”). **A3** Mean posterior pulmonary tuberculosis relative rate of change in case notification rates 1958-1963 vs. counterfactual (“slope effect”). **A2** ward specific and overall (red) “peak effect”, with 95% uncertainty interval. **B2** ward specific and overall (red) “level effect”, with 95% uncertainty interval. **A3** ward specific and overall (red) “slope effect”, with 95% uncertainty interval. UI: uncertainty interval.

There was an overall substantial decline in the pulmonary tuberculosis case notification rate in the year following in the ACF intervention (1958). Comparing 1958 to counterfactual trends, we estimate a 36% relative reduction in the Glasgow City case notification rate (relative rate: 0.64, 95% UI: 0.59 to 0.71), with consistent effects across wards (Figure 2). There was a positive correlation between the “peak effect” (increase in case notification rate in 1957 vs. counterfactual) and the “level effect” (decrease in case notification rate in 1958 vs. counterfactual), implying that greater increase in case detection in 1957 was associated with a smaller reduction in case notification in the immediate post-ACF year - Supplemental Figure S8.

In the post-ACF period (1958-1963), the rate of reduction in pulmonary tuberculosis case notification rates was greater than in the pre-ACF period. Overall, across Glasgow City during this six-year period, there was an annual rate of change of -5.4%, compared to -2.3% in the pre-ACF period. All wards had a markedly greater reduction in pulmonary tuberculosis case notification rates (“slope effect”) in the post-ACF period compared to the pre-ACF period (Figure 2). There was a positive correlation between greater “peak effect” of ACF, and lower relative reductions in the rate of change post-ACF (“slope effect”). There was no correlation between the “level effect” and the “slope effect” - Supplemental Figure S8.

In our extra-pulmonary tuberculosis model, the ACF intervention had little discernible peak (RR: 0.89, 95% UI: 0.72-1.09), level (RR: 0.85, 95% UI: 0.69-1.04), or slope effect (RR: 1.13, 95% UI: 0.76-1.64), either across the city, or at ward level (Supplemental Figures S9 and S10).

### Estimates of case detection

Assuming a 76% ACF coverage (as estimated from the total number screened and the estimated adult resident population), we estimated ward case detection ratios ranging from 60% to 92%, with a median (IQR) of 78% (72%-82%). Across wards, we found pre-ACF tuberculosis case notification rate and case detection ratios were positively correlated (corr=0.76), and case detection ratios were negatively correlated with ACF impact measured as pre/post notification ratio (corr= -0.55) – Supplemental Figure S11.

### Cases averted compared to counterfactual scenario

Across the City of Glasgow between 1958 and 1963, we estimate that, compared to the counterfactual scenario where the ACF intervention had not happened, there were 4,656 (95% UI: 3,670 to 5,725) pulmonary tuberculosis notifications averted, equivalent to a 43.1% (37.3% to 48.8%) reduction (Table 2).

**Table 2:** Impact of mass screening intervention in post-ACF period (1958-1963) compared to counterfactual scenario.

|  | Year | Difference in case notification rate (per 100,000 py, 95% UI) | Relative case notification rate (95% UI) | Tuberculosis notifications averted (95% UI) | Percentage difference in cases (95% UI) |
| --- | --- | --- | --- | --- | --- |
| PTB | 1958 | -64.8 (-79.4 to -50.9) | 0.64 (0.59 to 0.71) | 682.1 (535.2 to 835.2) | -35.5% (-41.0% to -29.5%) |
| PTB | 1959 | -69.7 (-84.9 to -55.3) | 0.61 (0.56 to 0.67) | 731.6 (580.6 to 891.5) | -38.9% (-44.2% to -33.2%) |
| PTB | 1960 | -74.0 (-90.5 to -58.6) | 0.58 (0.52 to 0.64) | 768.1 (608.0 to 939.9) | -42.0% (-47.5% to -36.3%) |
| PTB | 1961 | -77.8 (-96.0 to -61.3) | 0.55 (0.49 to 0.61) | 799.5 (629.3 to 985.9) | -45.0% (-50.8% to -39.0%) |
| PTB | 1962 | -81.3 (-100.9 to -63.3) | 0.52 (0.46 to 0.59) | 828.6 (645.2 to 1,029.0) | -47.9% (-54.0% to -41.4%) |
| PTB | 1963 | -84.3 (-105.8 to -64.9) | 0.49 (0.43 to 0.56) | 846.7 (651.5 to 1,063.0) | -50.6% (-57.1% to -43.7%) |
| **PTB** | **Overall (1958-1963)** |  |  | **4,656.5 (3,670.1 to 5,724.7)** | **-43.1% (-48.8% to -37.3%)** |
| EPTB | 1958 | -2.5 (-5.5 to 0.6) | 0.85 (0.69 to 1.04) | 0.7 (-0.2 to 2.0) | -14.9% (-31.2% to 4.1%) |
| EPTB | 1959 | -2.0 (-4.9 to 0.7) | 0.87 (0.71 to 1.05) | 0.6 (-0.2 to 1.8) | -13.2% (-29.1% to 5.2%) |
| EPTB | 1960 | -1.6 (-4.6 to 1.1) | 0.89 (0.71 to 1.09) | 0.5 (-0.3 to 1.7) | -11.4% (-29.5% to 9.5%) |
| EPTB | 1961 | -1.3 (-4.5 to 1.7) | 0.91 (0.69 to 1.17) | 0.4 (-0.5 to 1.6) | -9.3% (-31.4% to 16.6%) |
| **EPTB** | **Overall (1958-1963)** |  |  | **77.5 (-33.2 to 196.3)** | **-12.5% (-29.0% to 6.4%)** |

All estimates are compared to counterfactual scenario where active case finding intervention had not been implemented. PTB: Pulmonary tuberculosis; EPTB: Extra-pulmonary tuberculosis; py: person-years.

### Impact of active case finding by age group and sex

In the pre-intervention period, there were differences in pulmonary tuberculosis case notification trajectories by age group and sex, with in general declining rates in younger age groups, higher numbers of notifications among older men than women, and older men in particular having an upwards trend in notifications, compared to other groups (Supplemental Figure S12). With the intervention, there was a reduction in case notifications in young children (0-5 years) (Figure 3). The greatest difference between men and women in cases averted was in older age groups.

**Figure 3:**
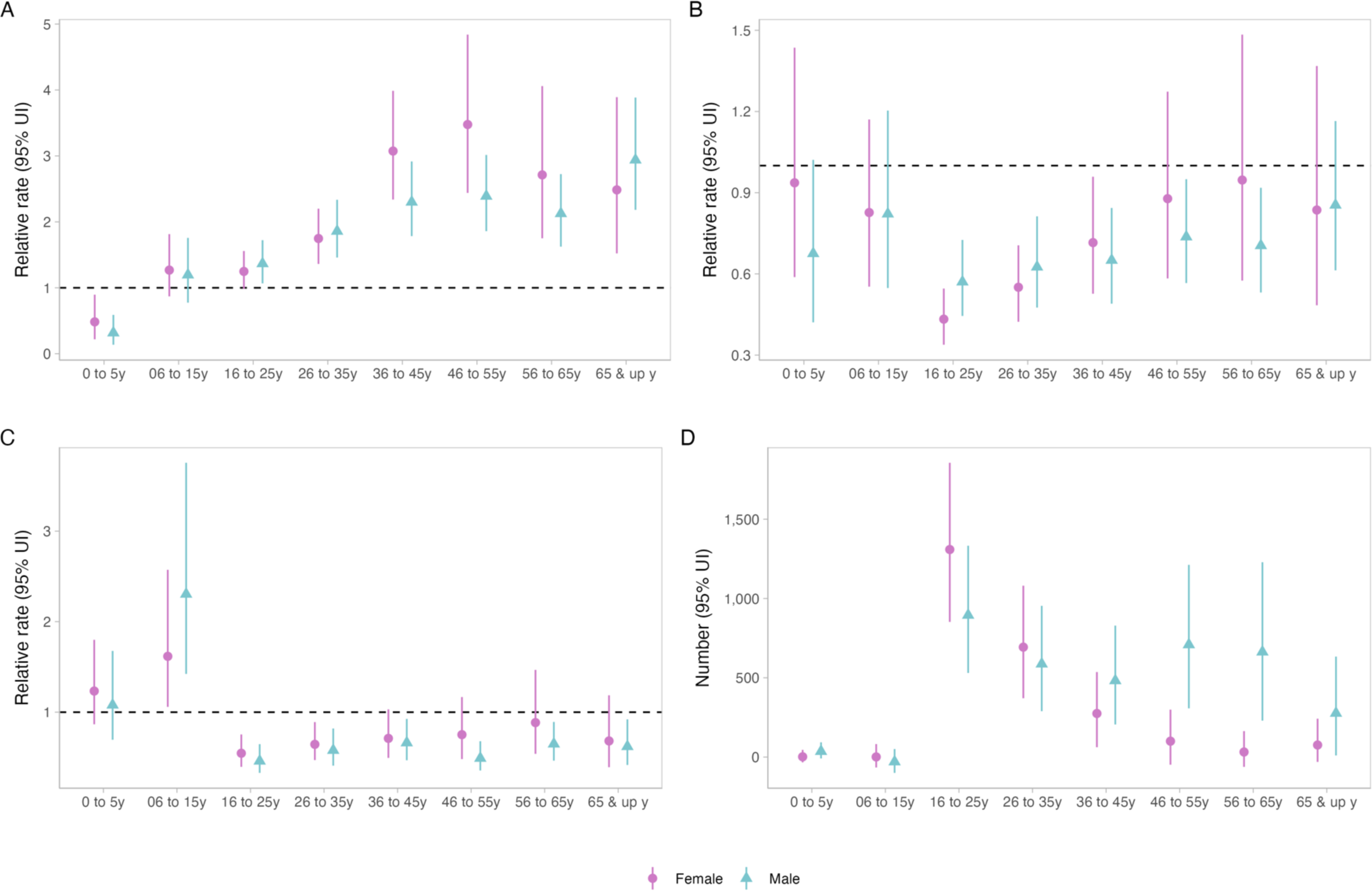
Impact of active case finding on case notifications by age group and sex, Glasgow City: 1950-1963. **A** “Peak effect” on case notifications (1957 vs. counterfactual). **B** “Level effect” on case notifications (1958 vs. counterfactual). **C** “Slope effect” on case notifications (1958-1963 vs. counterfactual). **D** Numbers of pulmonary tuberculosis notifications averted during 1958-1963, compared to counterfactual scenario. UI: Uncertainty interval.

## Discussion

The Glasgow mass miniature X-ray campaign for tuberculosis in 1957 was probably the largest single community-based active case finding intervention ever undertaken, with more than 715,000 people (622,349 adult Glasgow residents; 76% of the entire adult resident population) screened with miniature chest X-ray over just 5 weeks, equating to nearly 150,000 people screened per week of the campaign. In our analysis using modern epidemiological methods, we found that the mass screening intervention had a substantial immediate impact – doubling case notifications – and resulted in more rapid reductions in the citywide notification rate (5.4% per year) post intervention compared to the pre-intervention period (2.3%), with an estimated 4,656 notified pulmonary tuberculosis case notifications averted between 1958 and 1963 compared to the counterfactual scenario. Consistency in intervention effect across City wards gives confidence that the impact can be attributed to the intervention. By mostly omitting historical evidence of the impact of well-conducted mass X-ray screening programmes in global guideline development and contemporary discourse, it is likely that the potential impact of intensive, high-coverage ACF interventions to change epidemic trajectories has been under-estimated.

Neither systematic reviews of historical ACF interventions^14,22,23^ nor WHO guidelines for systematic tuberculosis screening^12,13^ have included these data from Glasgow in evidence synthesis. A review of historical mass miniature X-ray screening interventions from the 1930s to late 1960s identified 18 published reports from North America, Europe, and India^6^. Synthesising these data *Golub et al* concluded that mass miniature X-ray screening was highly effective at increasing case detection, particularly when programmes were well supported by community and health system engagement^6^. Likewise, *Miller et al* emphasised the importance of logistics at large scale to ACF programme success^24^. During the late 1950s however, several large cities in the UK – including Glasgow, Edinburgh, and Liverpool, as well as many cities and countries internationally (Brazil, Japan, Spain, Italy, Sweden) undertook mass miniature X-ray screening for tuberculosis. However, to the best of our knowledge evidence from these programmes have mostly not been collated, possibly because data reside only in municipal public health reports, rather than in more accessible scientific publications. Moreover, data included in both sets of WHO guidance for community-based systematic screening (2013^12^ and 2021^13^) include only a small number of historical studies. This represents a missed opportunity: many high tuberculosis burden countries have tuberculosis epidemiological characteristics similar to Europe and North American in the 1950s. By omitting historical evidence, there a danger that previous lessons learned are forgotten.

Only a small number of contemporary studies have investigated the impact of ACF for tuberculosis on case notification rates, with the large majority following up communities only in the period before and during the intervention^14^, and with nearly all showing large increases in case detection during implementation. In non-randomised studies, following up communities for prolonged periods after the ACF intervention is critical to identify effects that are temporary or short lived; our analysis here provides compelling evidence that a single intervention with high coverage can change epidemic trajectory. Two contemporary randomised trials of community-based active case finding (ACT3^15^ and TREATS^16^) have shown mixed results, with the more intensive ACT3 study in Vietnam resulting in a substantial reduction in prevalent pulmonary tuberculosis following universal sputum testing with Xpert, whereas no effect was identified from the less-intensive symptom-and chest X-ray based screening approach in TREATS. However, uptake of screening was low in these two trials, with only 45% of eligible participants screened by sputum Xpert in the best performing year of ACT3. We speculate that the very high coverage of chest X-ray screening achieved in Glasgow in 1957 was a major contributor to epidemiological impact, and potentially identified people with early and subclinical tuberculosis, rapidly reducing transmission.

Critically important to achieving high population coverage of tuberculosis screening was a programme of mass community engagement and mobilisation, supported by more than 12,000 volunteers. A recent qualitative synthesis of the community views of participating in ACF programmes emphasises that local ownership and leadership, ongoing support for people screened, and health systems strengthening to support the increased healthcare demands generated by mass screening are important determinants of intervention success^25^; The Glasgow mass screening campaign exemplified these principles, and we argue that community and health systems support within contemporary ACF programmes are often insufficient, and delivered in a “top-down” – rather than community-responsive – fashion, likely contributing to lower than anticipated participation and effectiveness.

The post-war period between 1948 and 1960 was a period of tremendous social change, and the 1957 Glasgow mass tuberculosis screening campaign needs to be contextualised alongside progressive improvements in living conditions, healthcare, and tuberculosis care and prevention. In Glasgow, “Slum clearances” commenced in the mid-1950s, particularly in wards in the centre and South of the city such as Camphill, Gorbals, Cowcaddens, and Govan, with new residential schemes established on the peripheries of the city. These changes in ward demographics can be seen in our supplementary figures. However, whilst the reduction in crowding and removal of dwellings in the worst condition is likely to have beneficial effects for reducing tuberculosis transmission, the slow improvements in social and housing conditions themselves are unlikely to account for the rapid and large changes in tuberculosis notifications that occurred immediately with and in the period after the mass screening campaign. Future studies may attempt to investigate associations between the numbers of dwellings demolished in slum clearances, change in other markers of social deprivation, and magnitude of impact of the intervention.

We found strong evidence that the impact of the ACF intervention differed by age group and sex. In young children (<5 years, who themselves weren’t eligible for screening, although a small number of children <15 years did undergo chest X-ray), case notification rates decreased during the intervention year in contrast to other age groups which saw large increases; we speculate that this may indicate evidence of ACF shortening infectious duration and providing early beneficial impact on transmission. For adults, contemporary data from high tuberculosis burden countries show that men have twice the prevalence of undiagnosed pulmonary tuberculosis than women^26^. Men are less likely to participate in prevalence surveys in Africa^27^ and Asia^28^, and in active case finding trials^15,16,29^, prompting calls to target interventions towards men^17^. However, if tuberculosis prevalence was indeed higher in men than women in Glasgow in the 1950s, our analysis suggests that rapid delivery of high coverage of high sensitivity screening (with chest X-ray) – rather than interventions targeted specifically at men – may achieve benefits for all groups, but especially those where disease burden is likely to have been highest. We found no effect on extrapulmonary tuberculosis; this is to be expected, as extrapulmonary tuberculosis notification rates were already low by the time the intervention started, and mass chest X-ray screening targets pulmonary tuberculosis. Importantly, our analysis could only investigate population-level effects on case notifications; previous reviews have emphasised the dearth of information on the individual-level benefits and harms of participating in community tuberculosis screening^30^. There was little description available of microbiological testing results or treatment outcomes, and it is possible that there was over-treatment of tuberculosis in people screened, or indeed treatment of people with very early subclinical TB which would not have usually been detected and treated. Future ACF trials should systematically record individual-level benefits and harms.

Despite the high-quality data available and the rigorous epidemiological methods used, there are some limitations to this analysis. We assumed a linear trend in tuberculosis case notification rates in the pre-ACF period, and projected this forward for the counterfactual scenario in the post-ACF period. It is possible however that – in the absence of the intervention – the rate of decline may have accelerated downwards in the late 1950s to early 1960s, leading us to over-estimate impact. We relied on available census data and official ward population estimates from government and municipal sources; however, the population of Glasgow changed rapidly in the late 1950s, and these may have over-or under-estimated population denominators, particularly for some wards. We did not have comprehensive data on microbiological results or treatment outcomes for notified cases throughout the study period, nor for participants in the intervention; a post-intervention study of prevalence was not done. Finally, while the rapidity, magnitude, and consistency of impact across wards strongly support of a direct causal effect, other factors (“slum clearances”, mobility, general improvements in living conditions, access to healthcare and air quality) may have contributed to improvements in tuberculosis epidemiology.

In conclusion, rapid mass miniature chest X-ray screening of over 715,000 people in Glasgow, Scotland (76% of the entire adult city population within 5 weeks) resulted in an accelerated and sustained reduction in tuberculosis case notification rates in Glasgow, with an estimated 4,656 pulmonary tuberculosis case notifications averted in the six years following the intervention, and potentially evidence of an intervention effect on reducing transmission to young children. Previous attempts to synthesise data from tuberculosis mass screening programmes have mostly ignored examples from the 1950-1960s held in municipal health departments, with the consequence that our understanding of the population benefit of ACF has likely been underestimated. Cities in today’s high tuberculosis burden countries may benefit from mass chest X-ray screening supported by high levels of health system and community engagement, alongside improvements in living conditions.

## Data Availability

Data and code to reproduce analysis are available at https://github.com/petermacp/glasgow-cxr.

## Acknowledgements

We gratefully acknowledge the staff of the Special Collections at the City of Glasgow Archives for facilitating access to historical reports and maps. We also acknowledge Callan T MacPherson and Isabella L MacPherson for assistance in data extraction from historical records.

## Supplemental Material

**Supplemental Figure S1:**
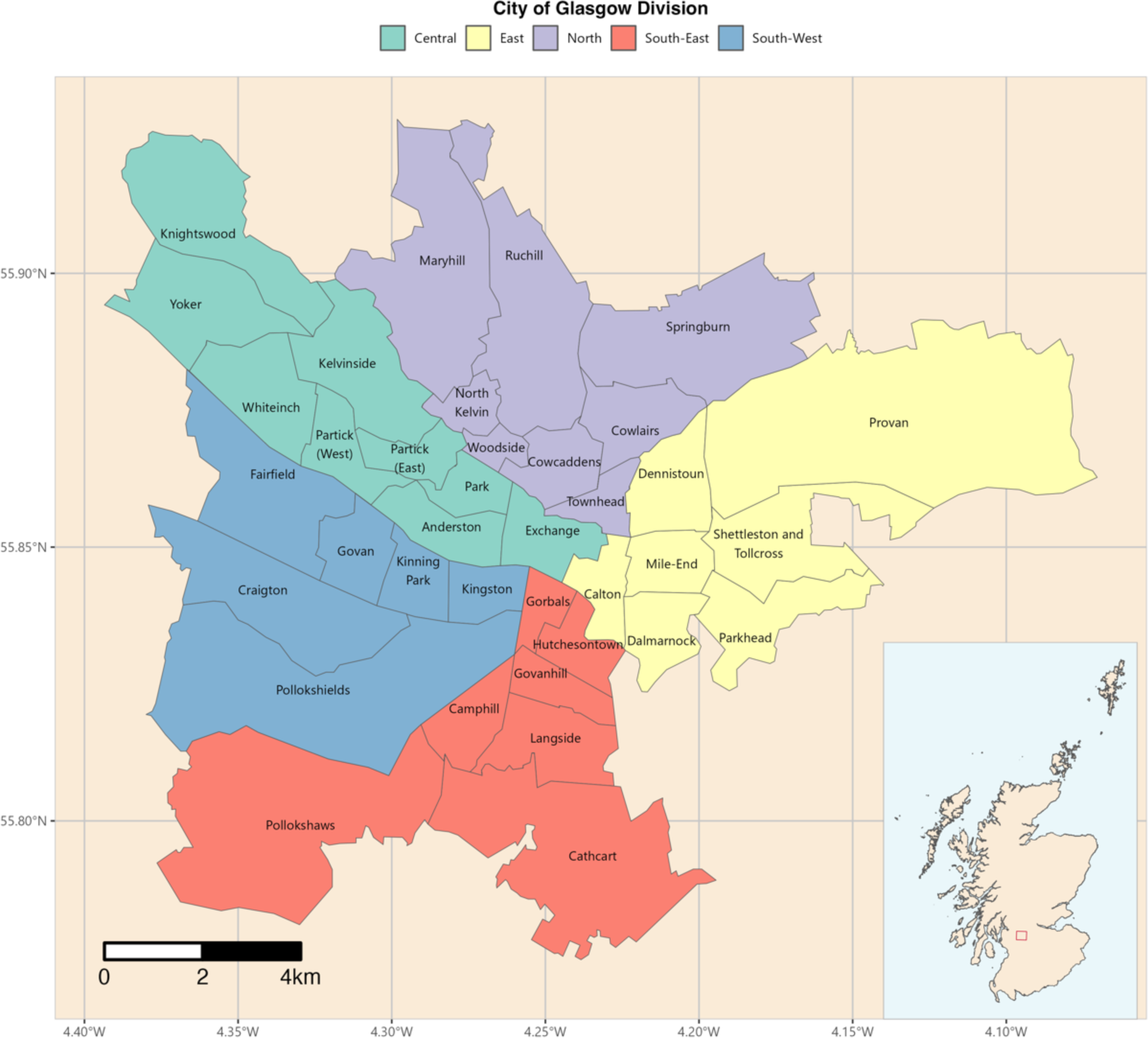
Divisions and Wards of the City of Glasgow, 1951. Red box in inset map of Scotland shows location of the main figure (City of Glasgow). Ward boundaries obtained from a scale 1951-1952 Post Office Directory map drawn by John Bartholomew FRSG held within the City of Glasgow Archive Special Collections (item PSI-52), digitalised using QGIS 3.34.1.

## Supplemental equation 1

Annual tuberculosis case notification rates are estimated using the following model:

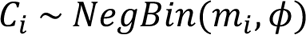

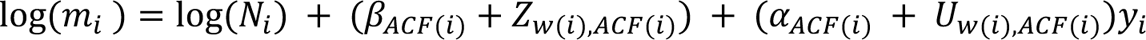

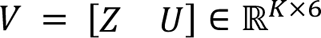

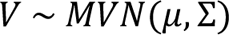

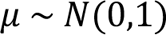

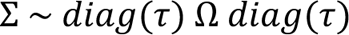

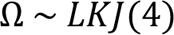

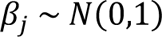

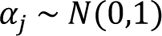

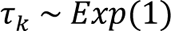

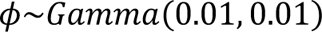

Where i ranges over 1,…,N indexing each row/datapoint; w() maps each row onto its ward (of which there are K); ACF() maps each row onto its time period: one of ‘pre’, ‘during’, ‘post’

## Supplemental Methods

### Estimates of incidence and case detection

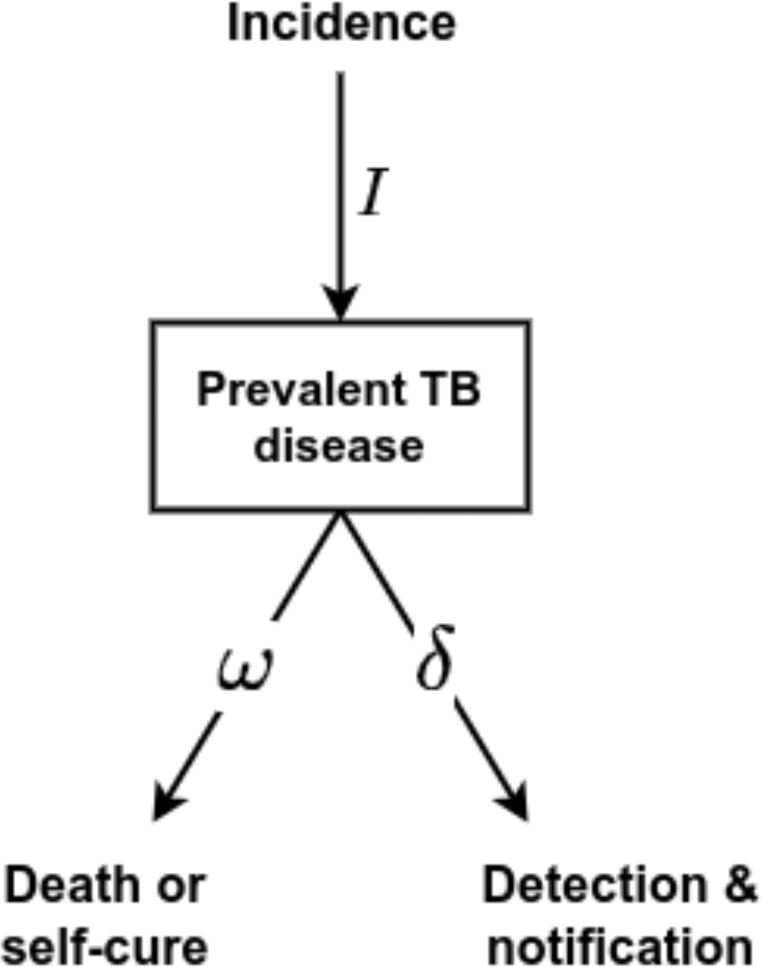

We used a simple competing hazard model of TB detection. Incident TB disease is the inflow to the prevalent pool; competing hazards of detection/notification (*δ*) and deaths/self-cure (*ω*) apply to the prevalent pool. The inverse of the rate of death/self-cure represents the mean duration of TB in the absence of treatment:

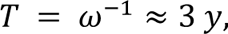

the estimate of a 3 years duration based on Tiemersma et al.

Under an equilibrium assumption, we can write down formulae for the case detection ratio (CDR), and in terms of prevalence (*P*) and notifications (*N*):

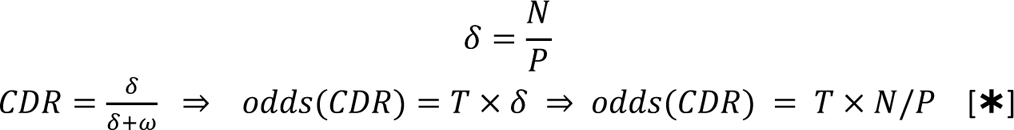

Under the assumption that the increase in notifications (*N*_*acf*_ − *N*_*pre*_) from just before (*N*_*pre*_) to during ACF (*N*_*acf*_) represents a coverage fraction ACF of the prevalent pool being found:

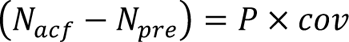

We can use this together with equation for *CDR* above to find

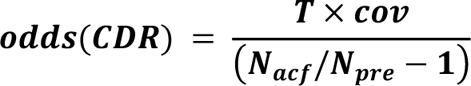

The incidence prior to ACF (*I*_*pre*_) can then be estimated as:

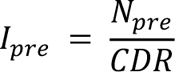

---

**Supplemental Figure S2:**
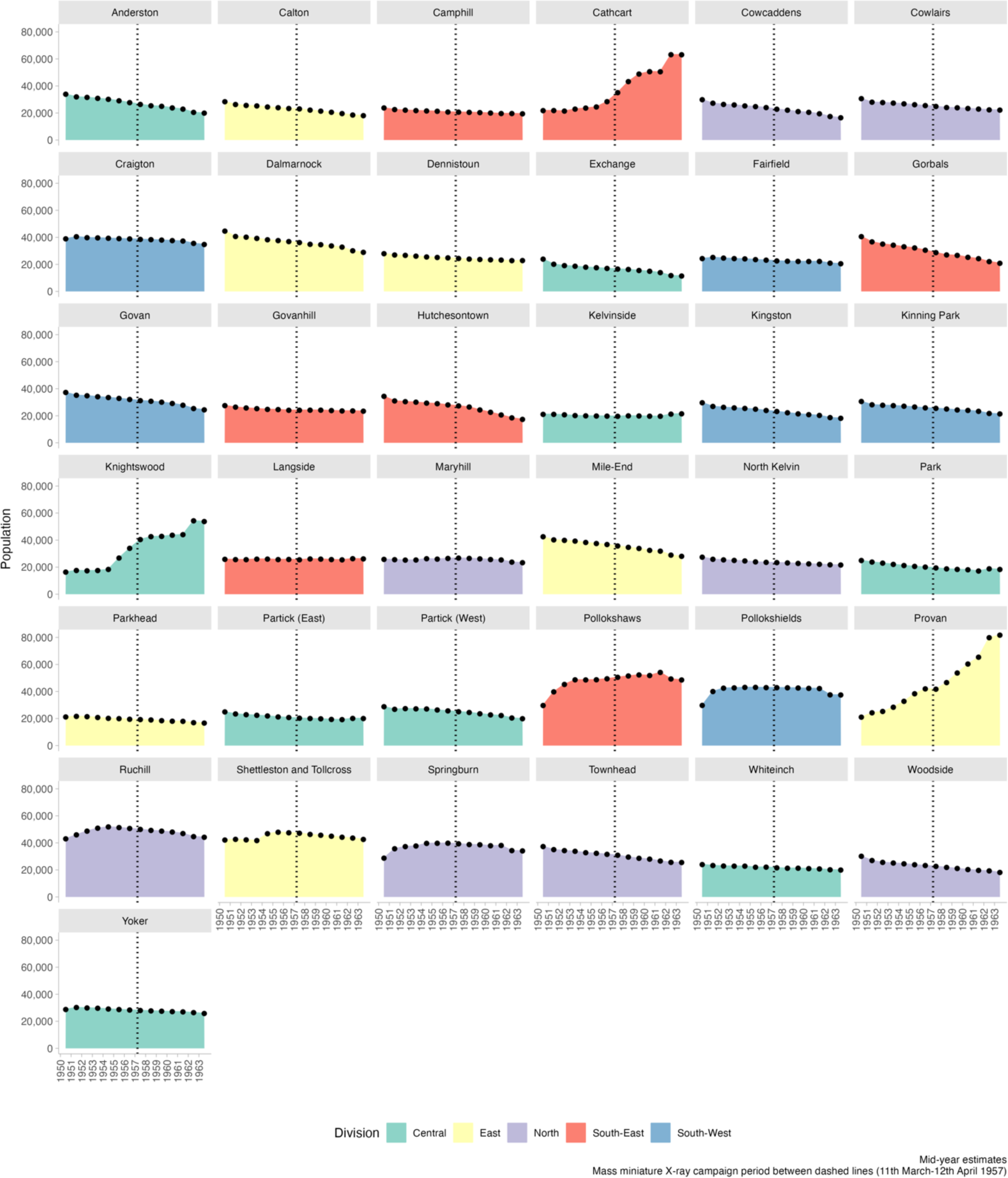
Glasgow City population by ward.

**Supplemental Figure S3:**
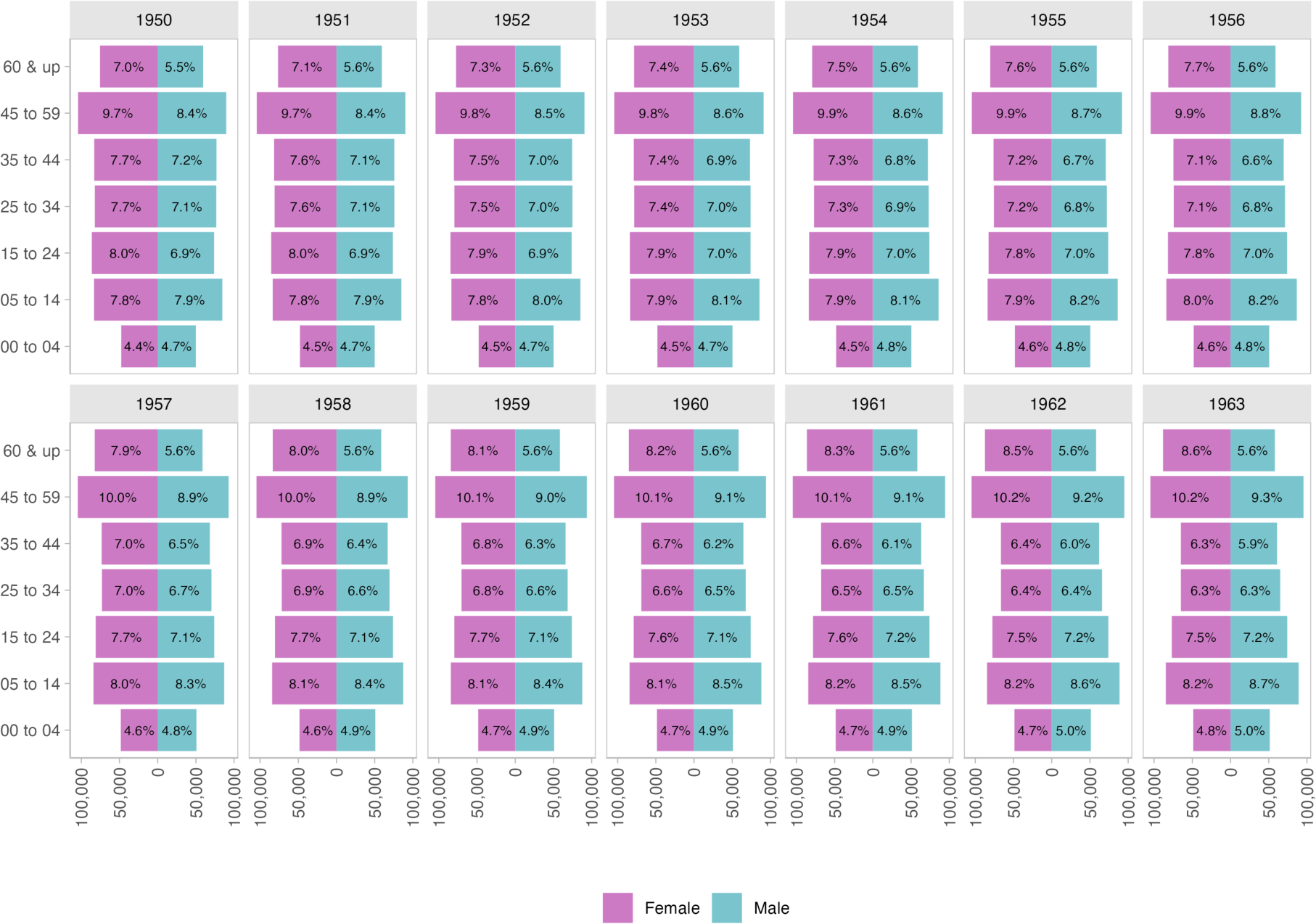
Glasgow City population pyramids by year.

**Supplemental Figure S4:**
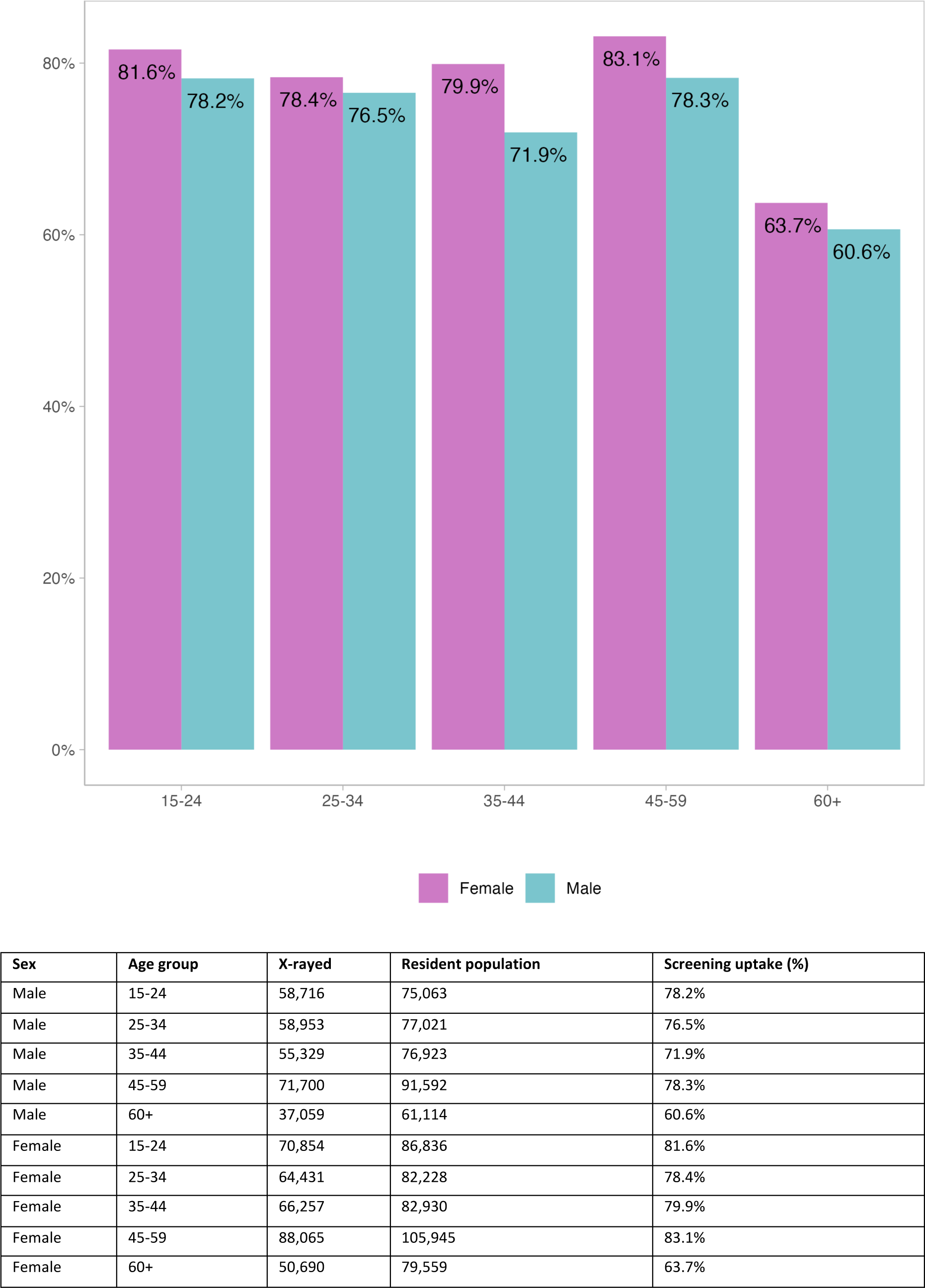
Uptake of tuberculosis screening by age and sex.

**Supplemental Figure S5:**
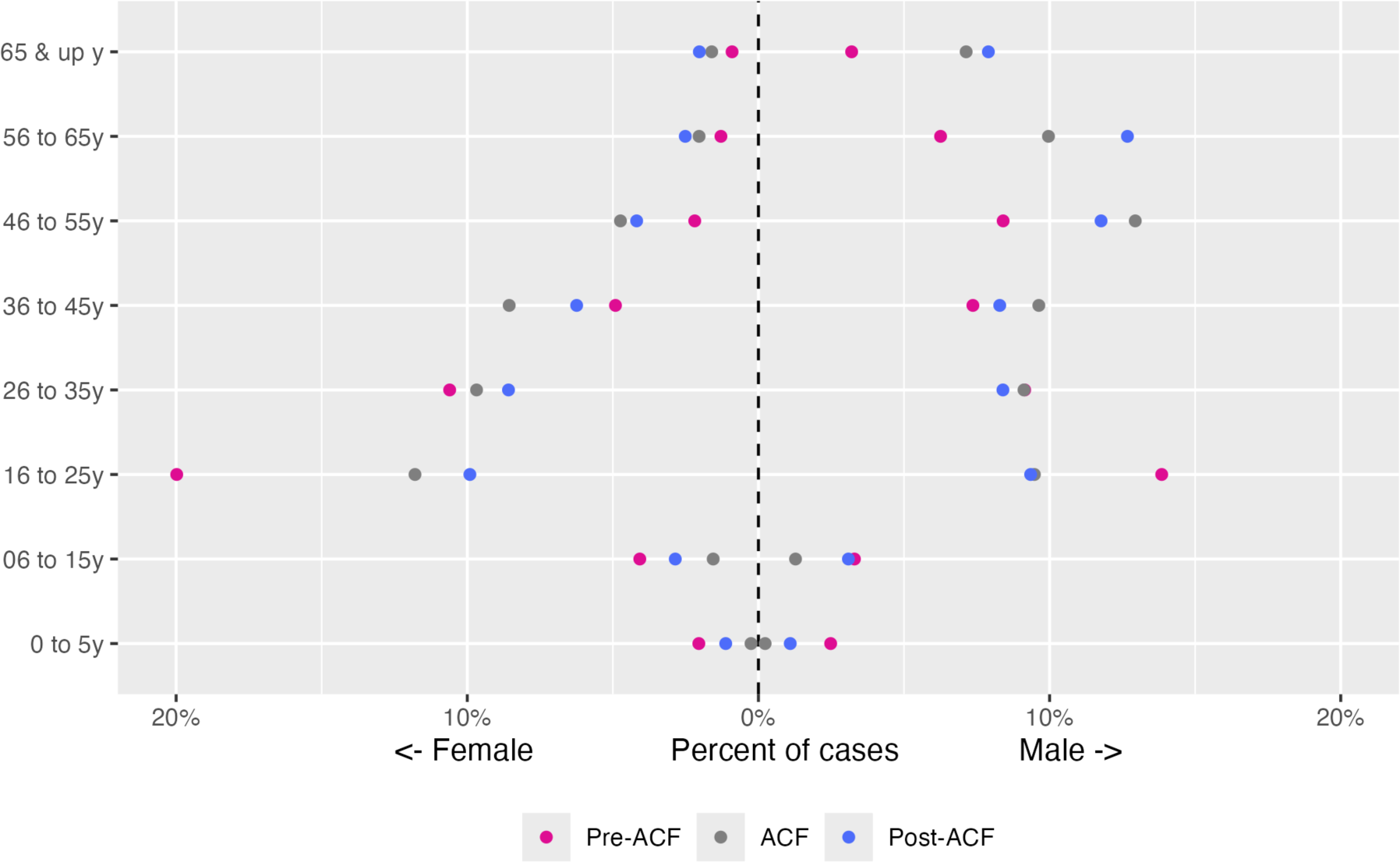
Distribution of pulmonary tuberculosis cases by age group and sex in Glasgow by study period. ACF: active case finding

**Supplemental Figure S6:**
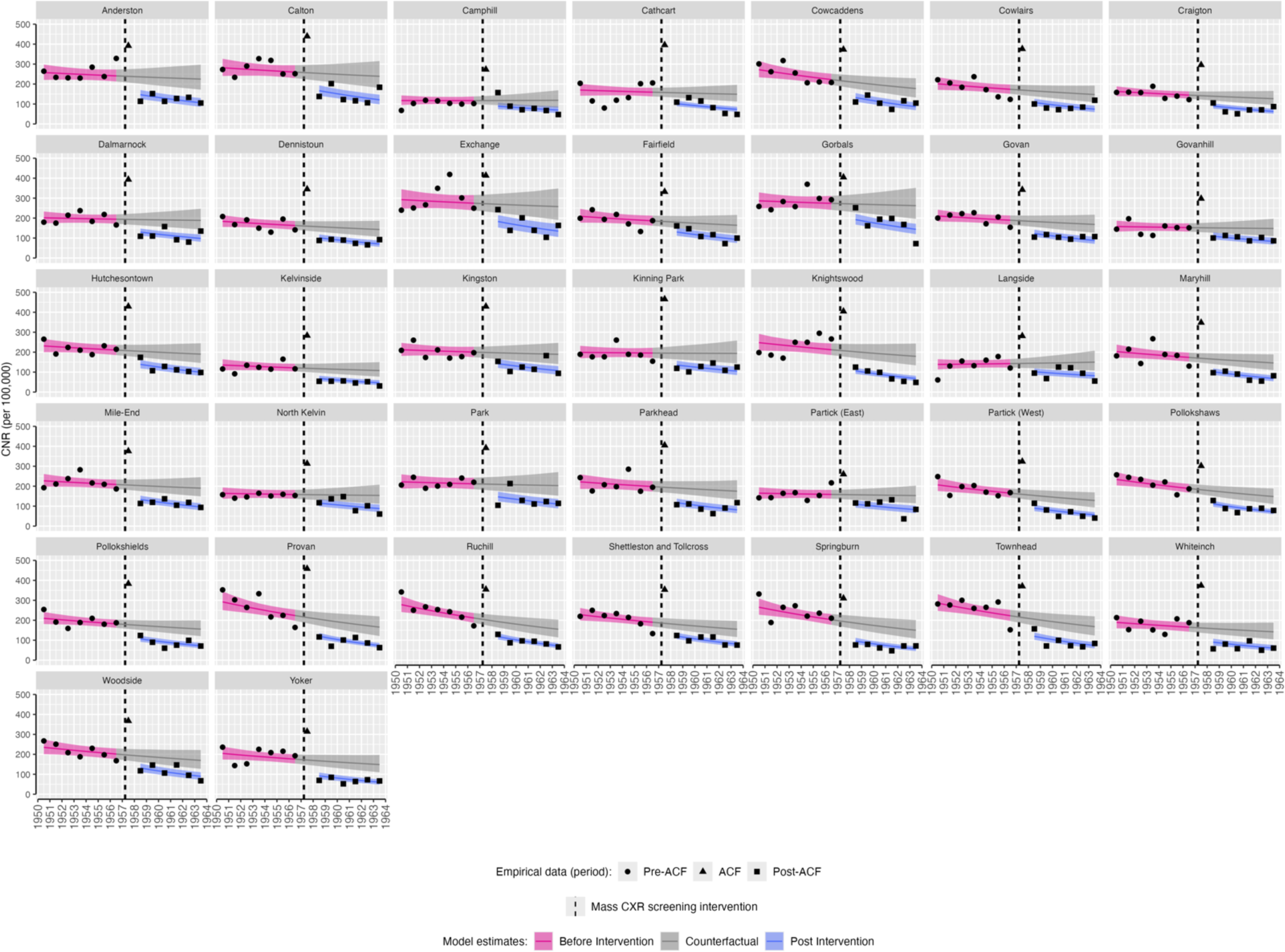
Pulmonary tuberculosis case notification rates by Glasgow Ward, 1950-1963. Empirical and modelled case notification rates (per 100,000 population) by ward, with counterfactual of no active case finding intervention. The mass miniature X-ray active case finding campaign occurred between dashed lines (11th March-12th April 1957). CNR: case notification rate. ACF: active case finding.

**Supplemental Figure S7:**
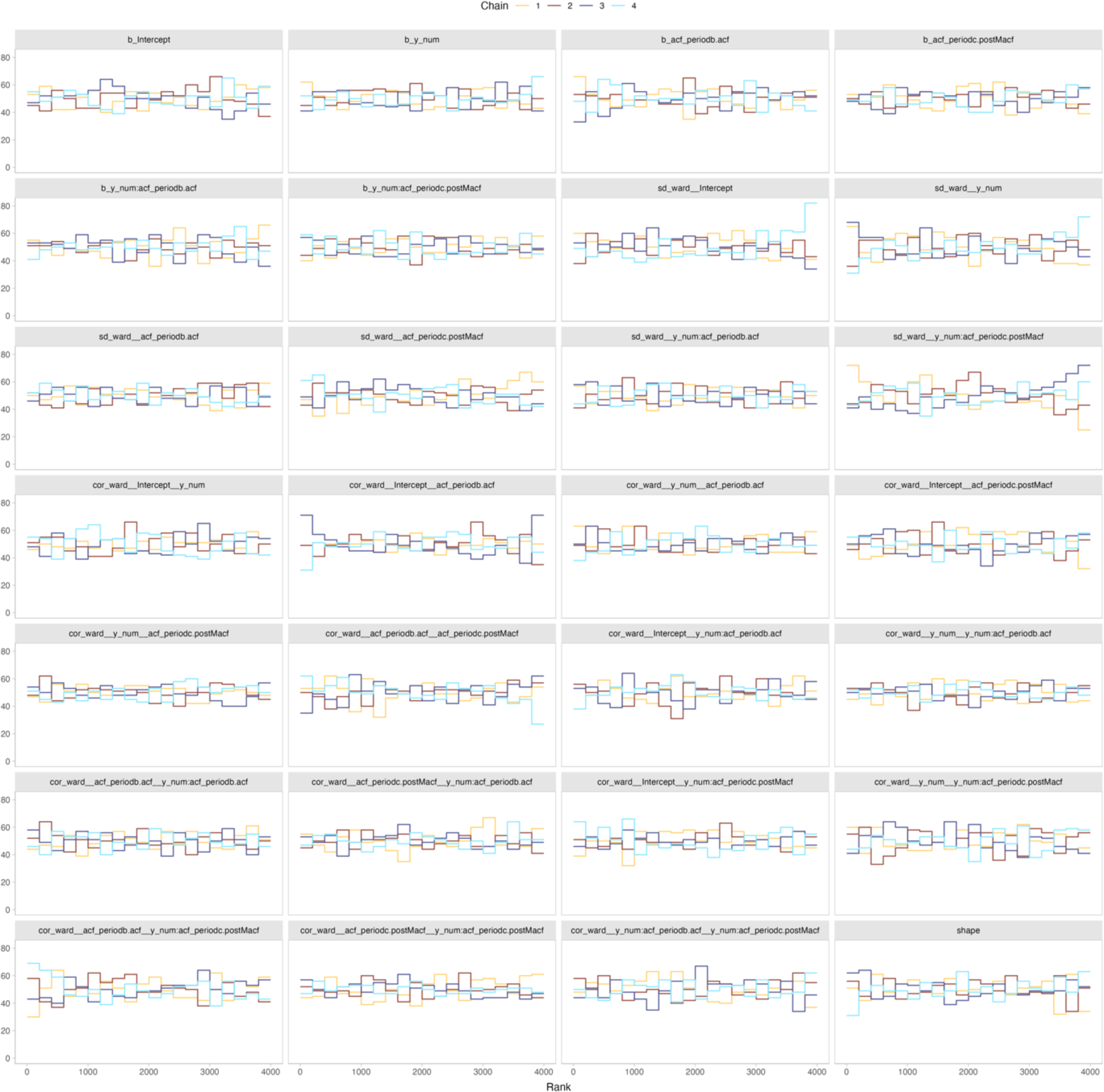
Rank plots of Markov chain Monte Carlo draws from pulmonary tuberculosis model.

**Supplemental Table S1:**
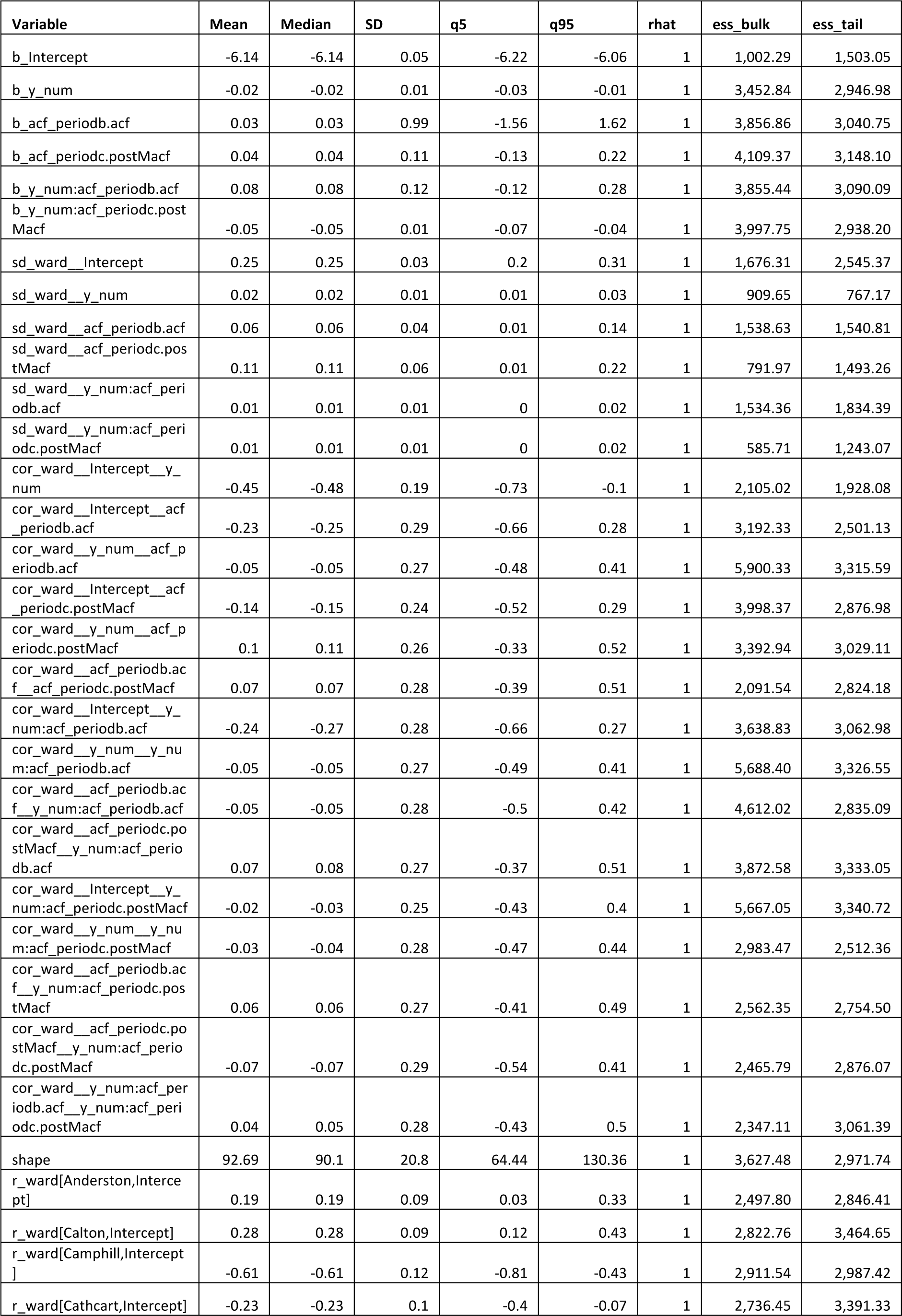
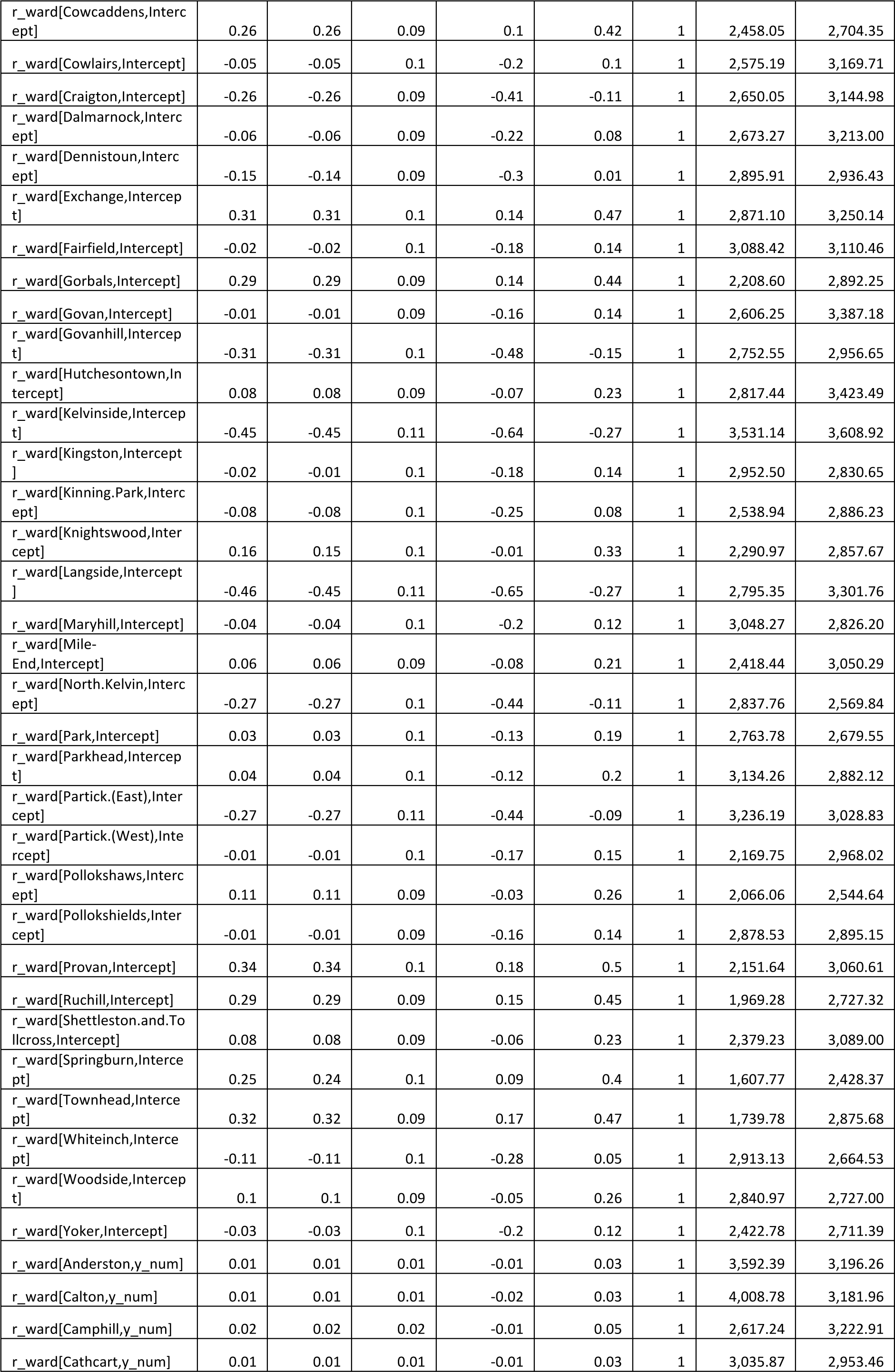
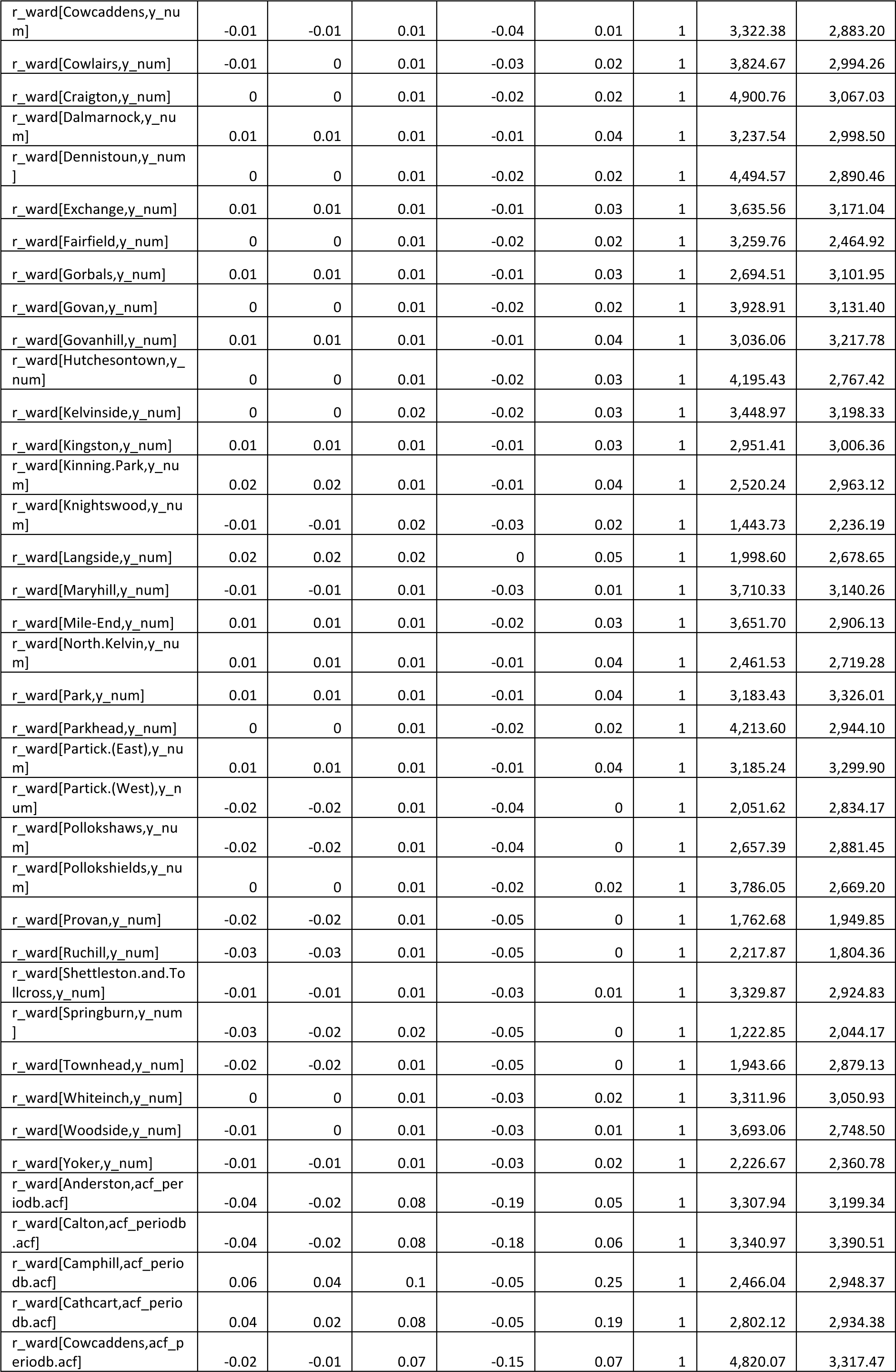
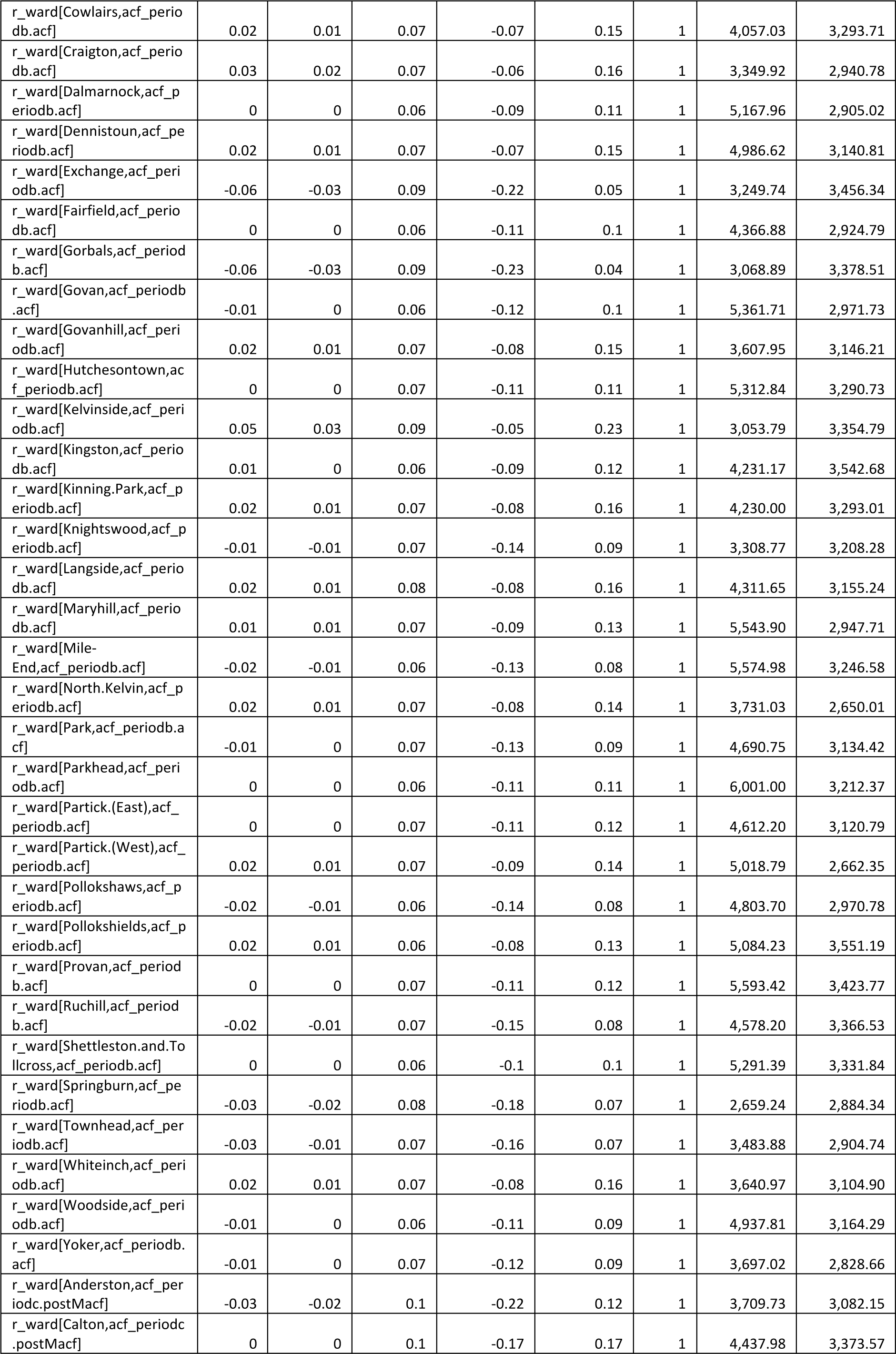
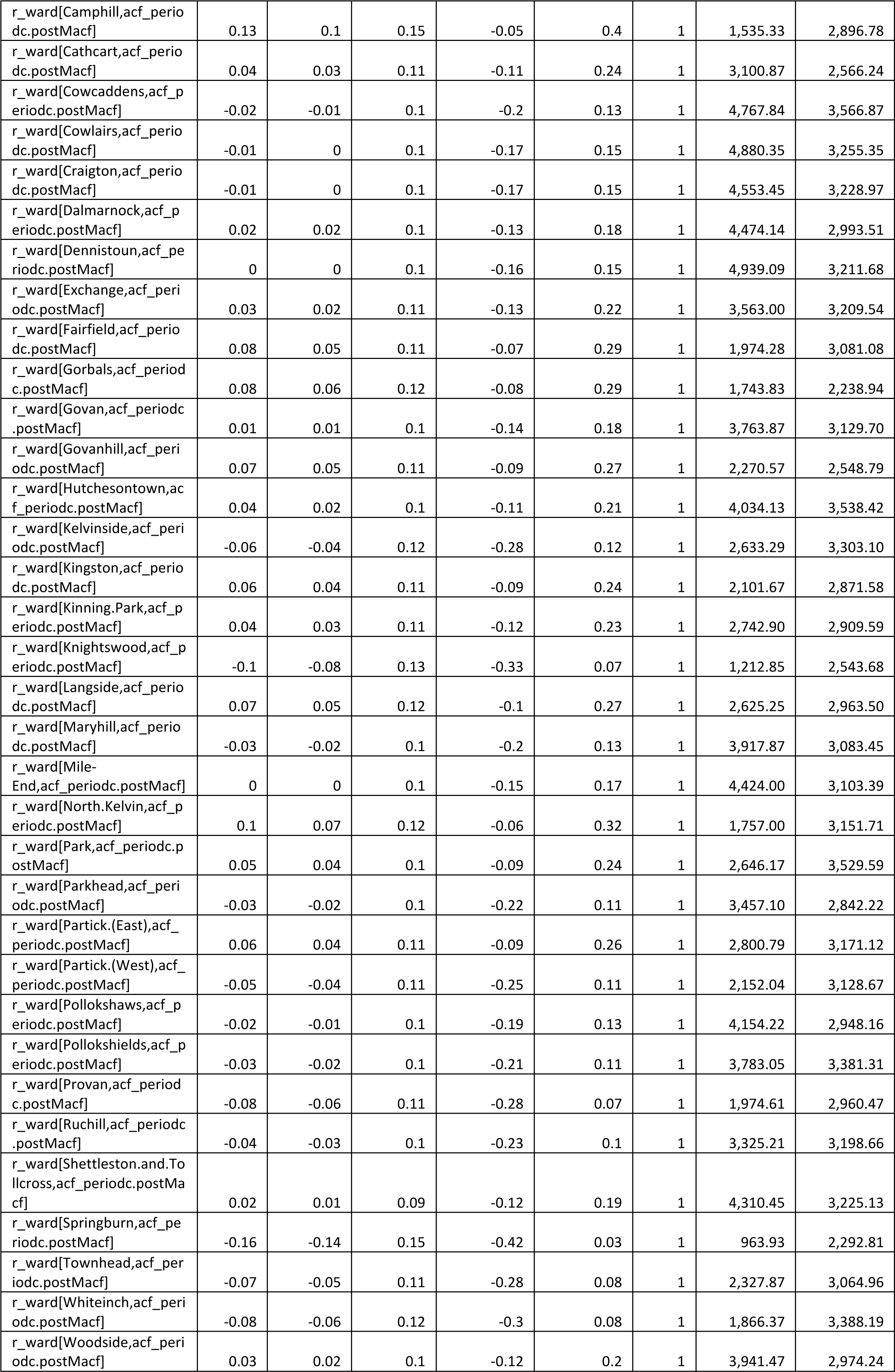
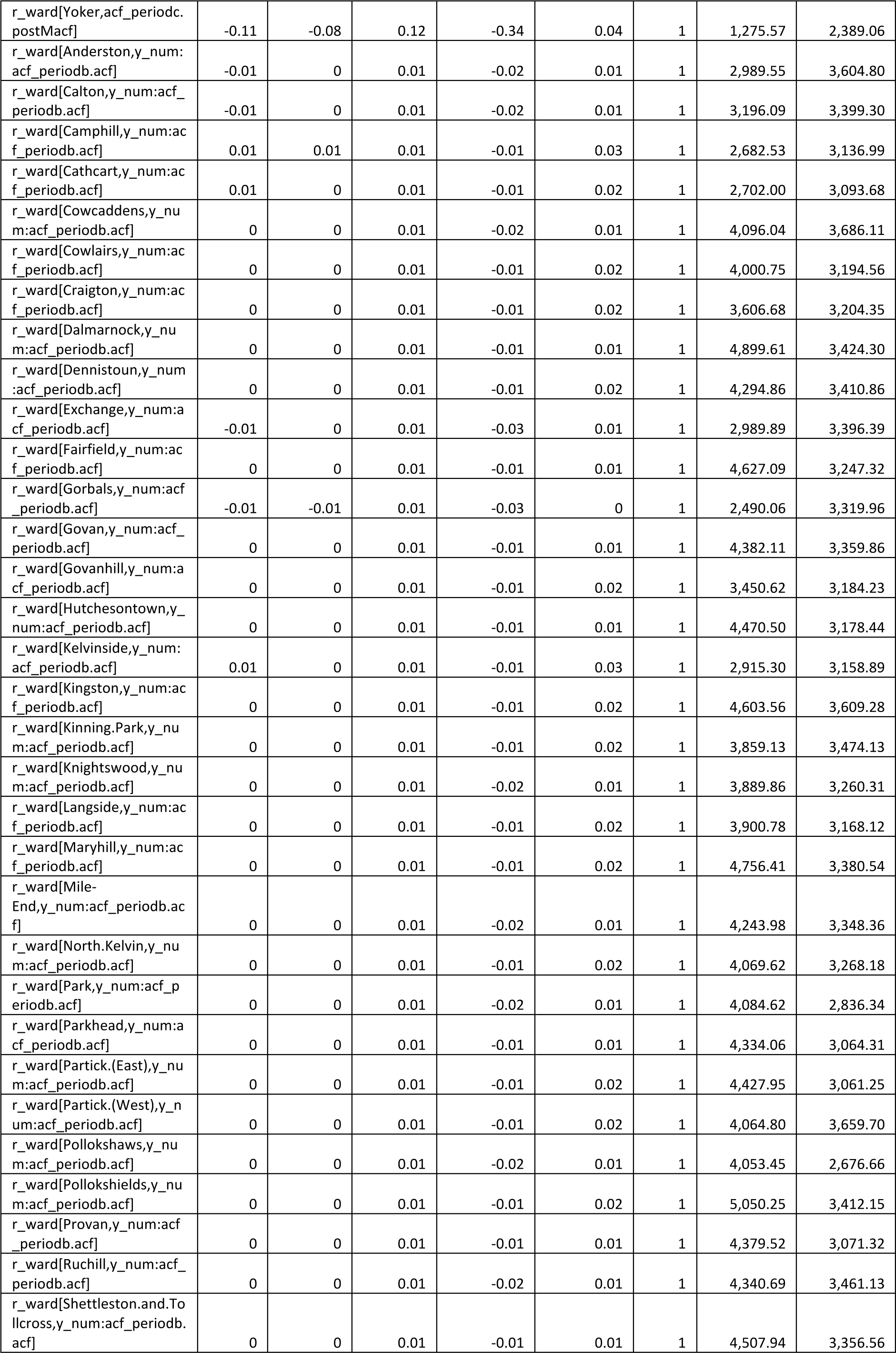
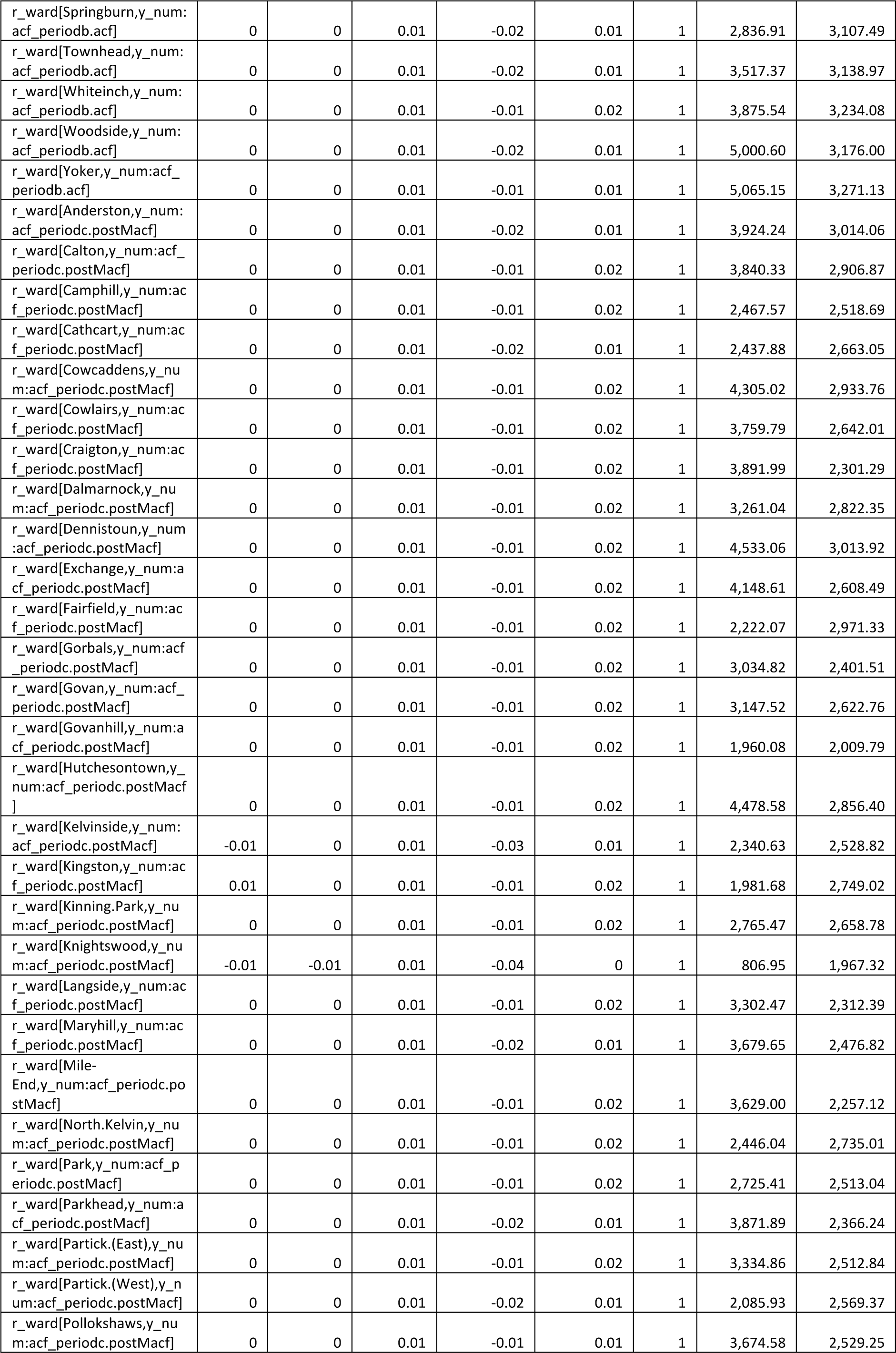
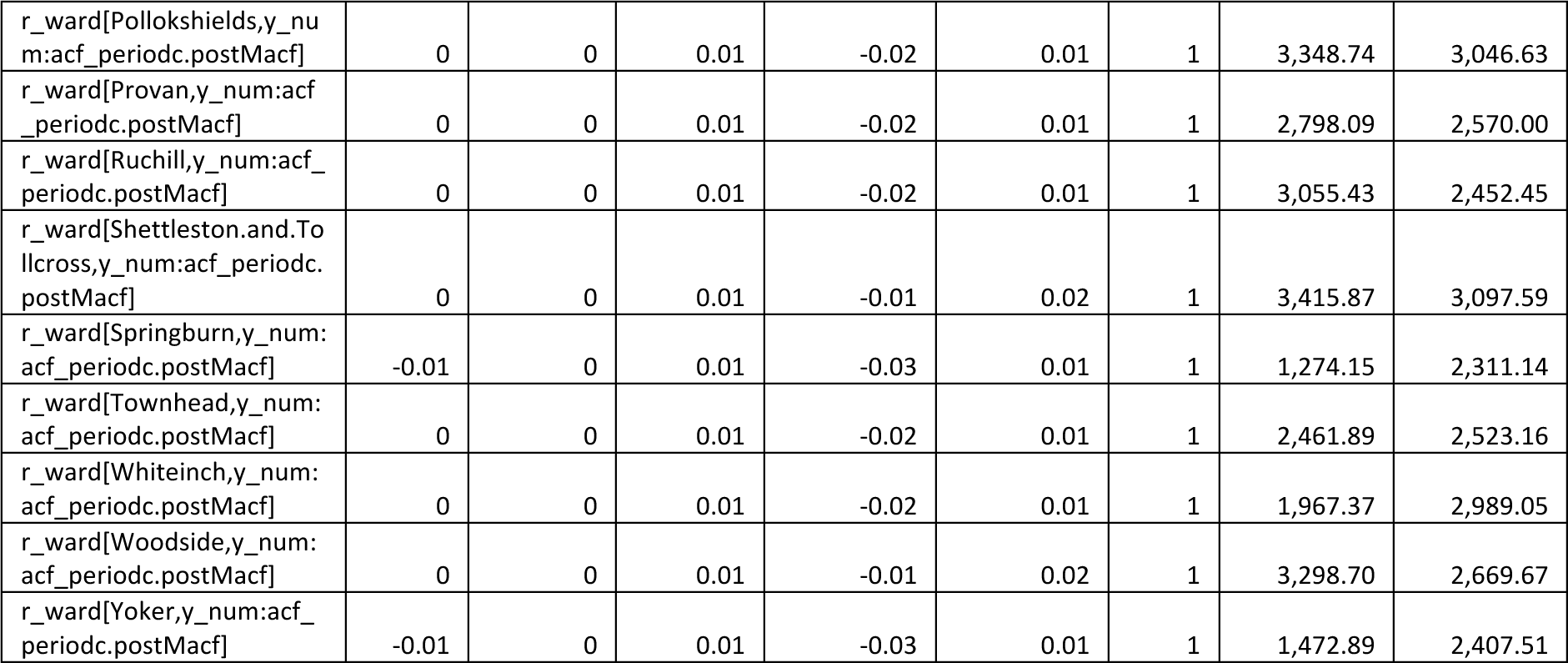
Summary of posterior draws from pulmonary TB mode.

**Supplementary Figure S8:**
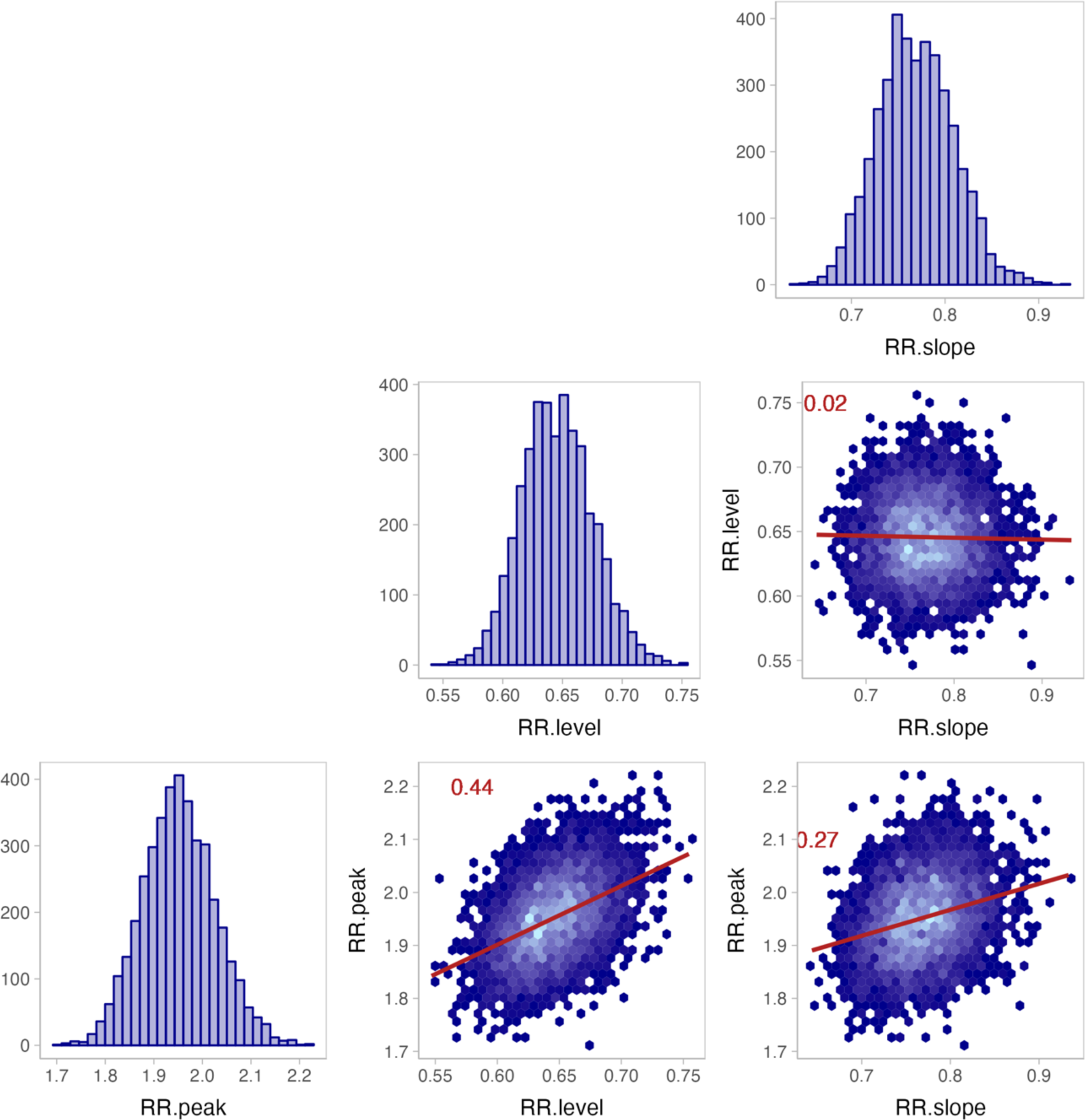
Posterior distributions of, and correlations between “peak effect”, “level effect”, and “slope effect” of active case finding intervention.

**Supplemental Figure S9:**
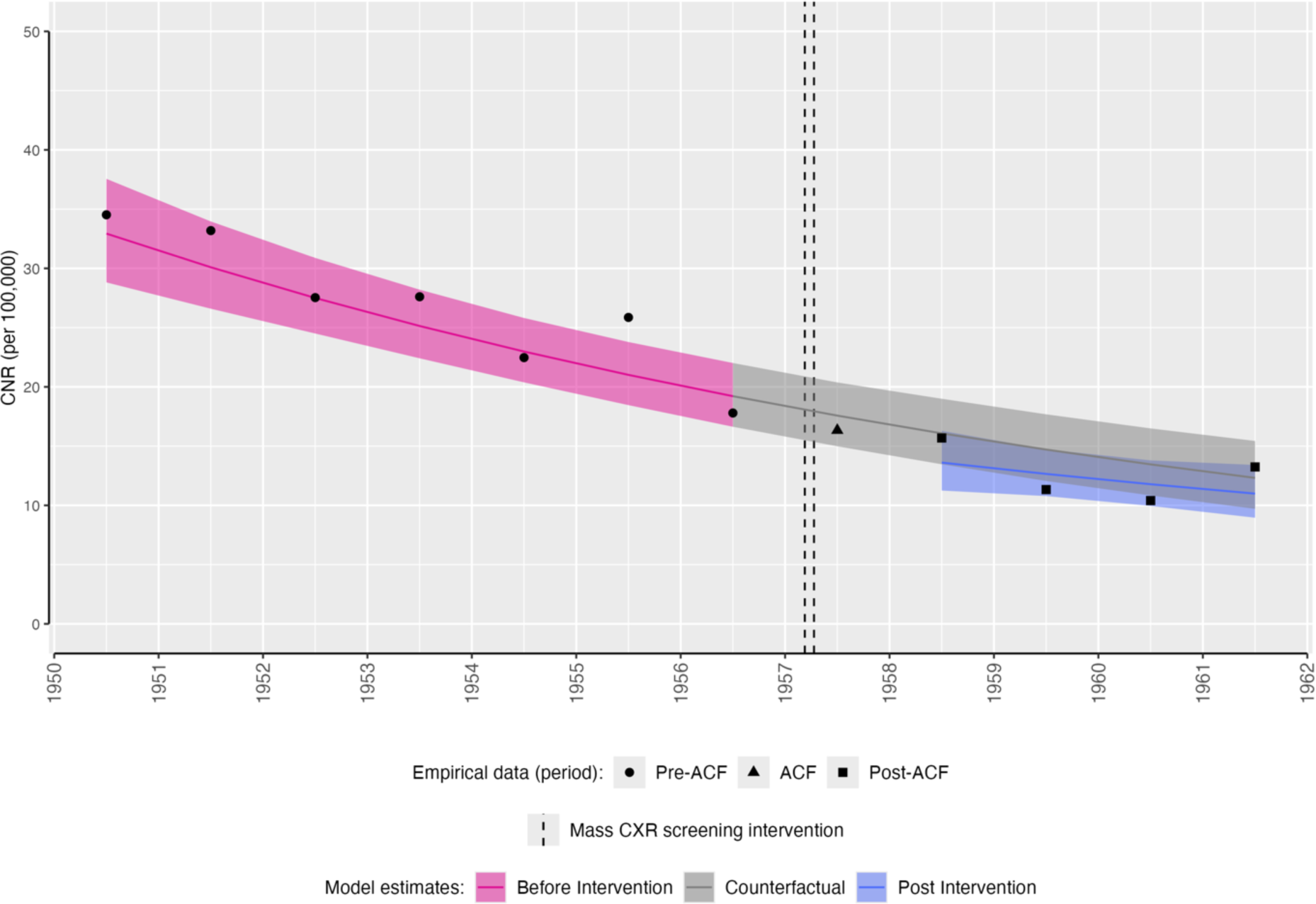
Impact of active case finding intervention on extra-pulmonary tuberculosis case notification rates, Glasgow City. Empirical and modelled case notification rates (per 100,000 population) by ward, with counterfactual of no active case finding intervention. The mass miniature X-ray active case finding campaign occurred between dashed lines (11th March-12th April 1957). CNR: case notification rate. ACF: active case finding.

**Supplemental Figure S10:**
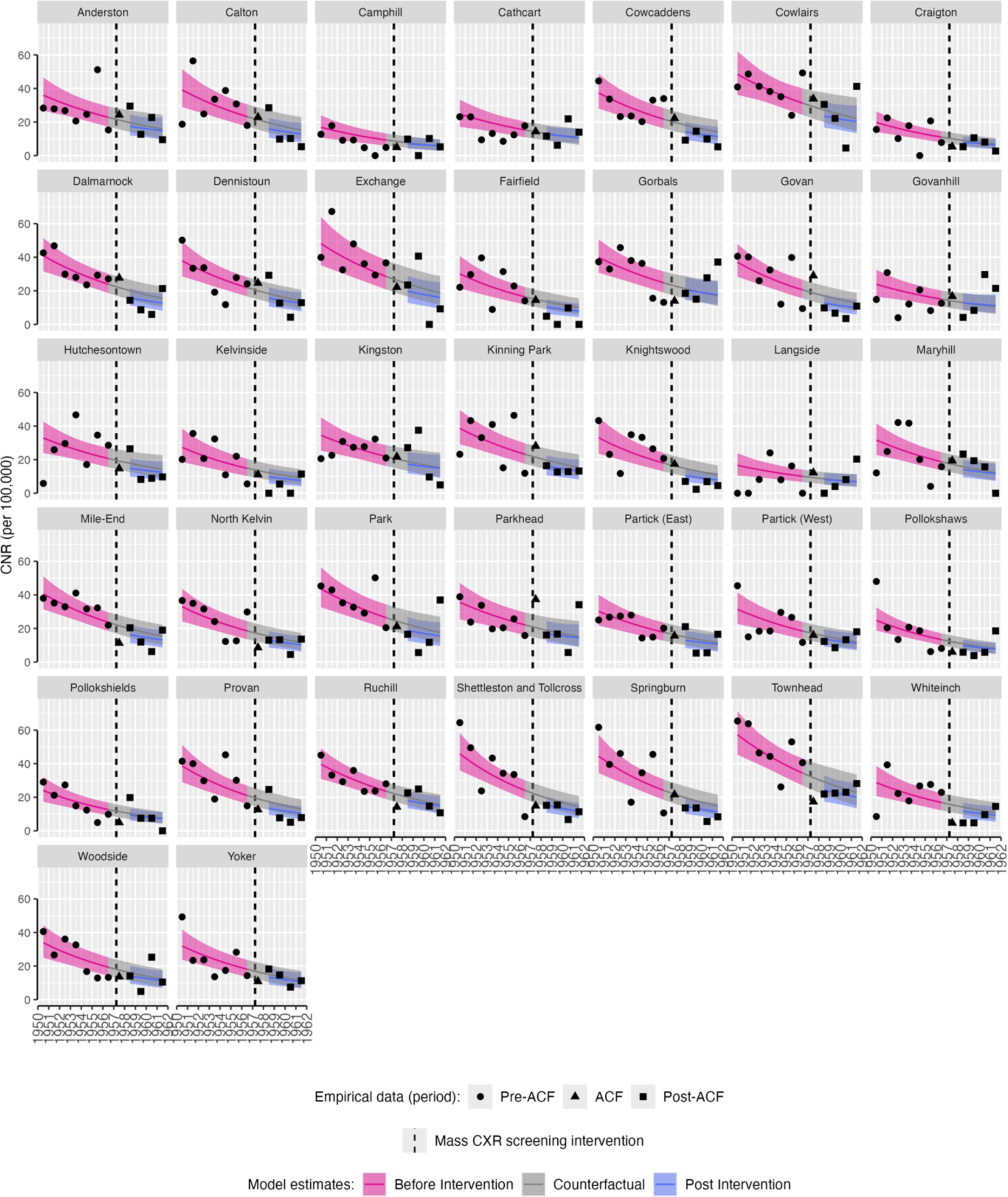
Impact of active case finding intervention on extra-pulmonary tuberculosis case notification rates, ward-level. Empirical and modelled case notification rates (per 100,000 population) by ward, with counterfactual of no active case finding intervention. The mass miniature X-ray active case finding campaign occurred between dashed lines (11th March-12th April 1957). CNR: case notification rate. ACF: active case finding.

**Supplemental Figure S11:**
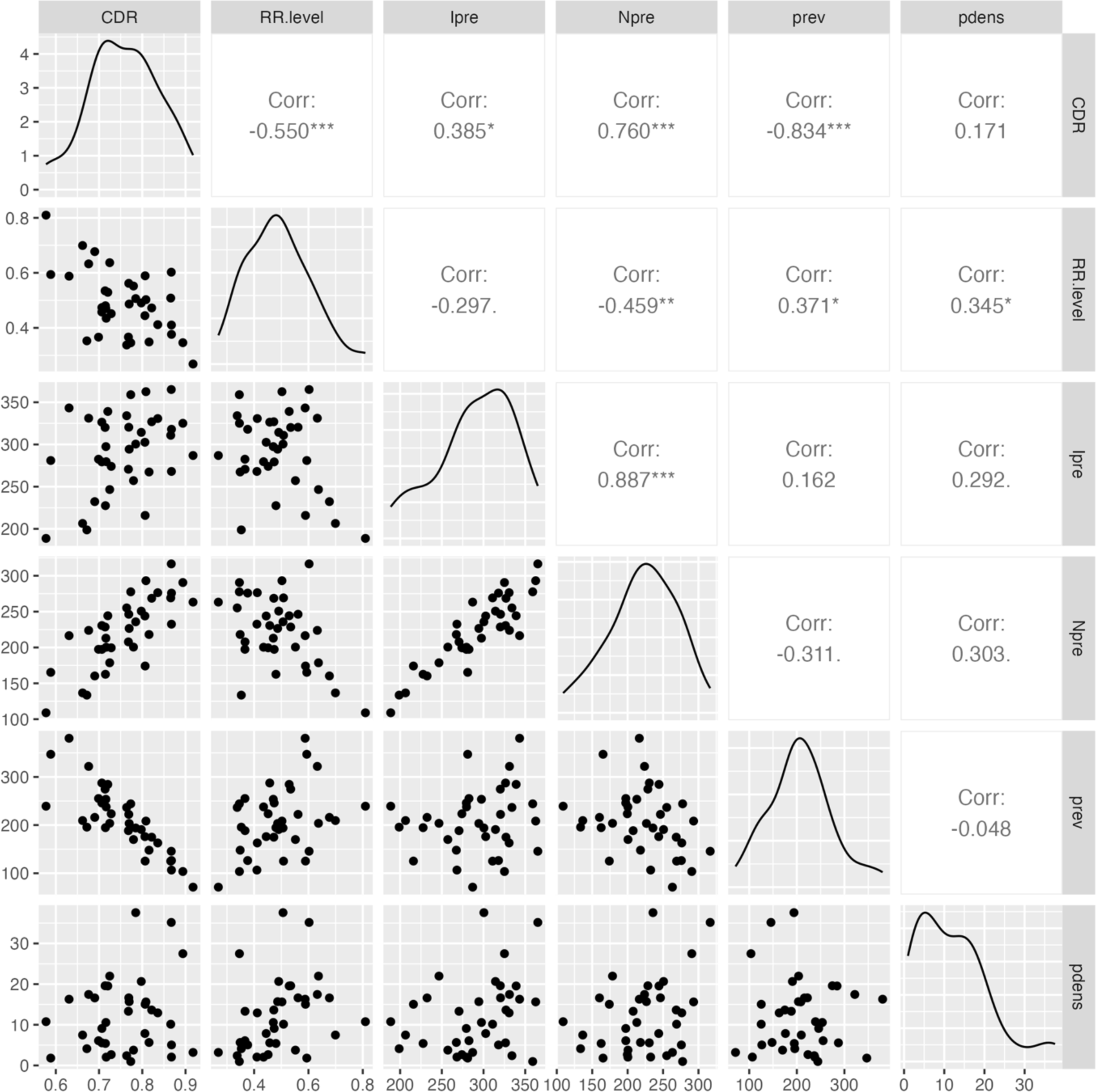
Correlation between estimated case detection active case finding impact. Points are Ward specific values. CDR: Case detection rate; RR.level: Mean posterior relative pulmonary TB case notification rate in 1958 vs. counterfactual (“level effect”); Ipre: mean estimated incidence per 100,000 in pre-ACF period; Npre: mean case notification rate per 100,000 in pre-ACF period; prev: estimated prevalence per 100,000; pdens: population density (1000 people per square kilometre).

**Supplemental Figure S12:**
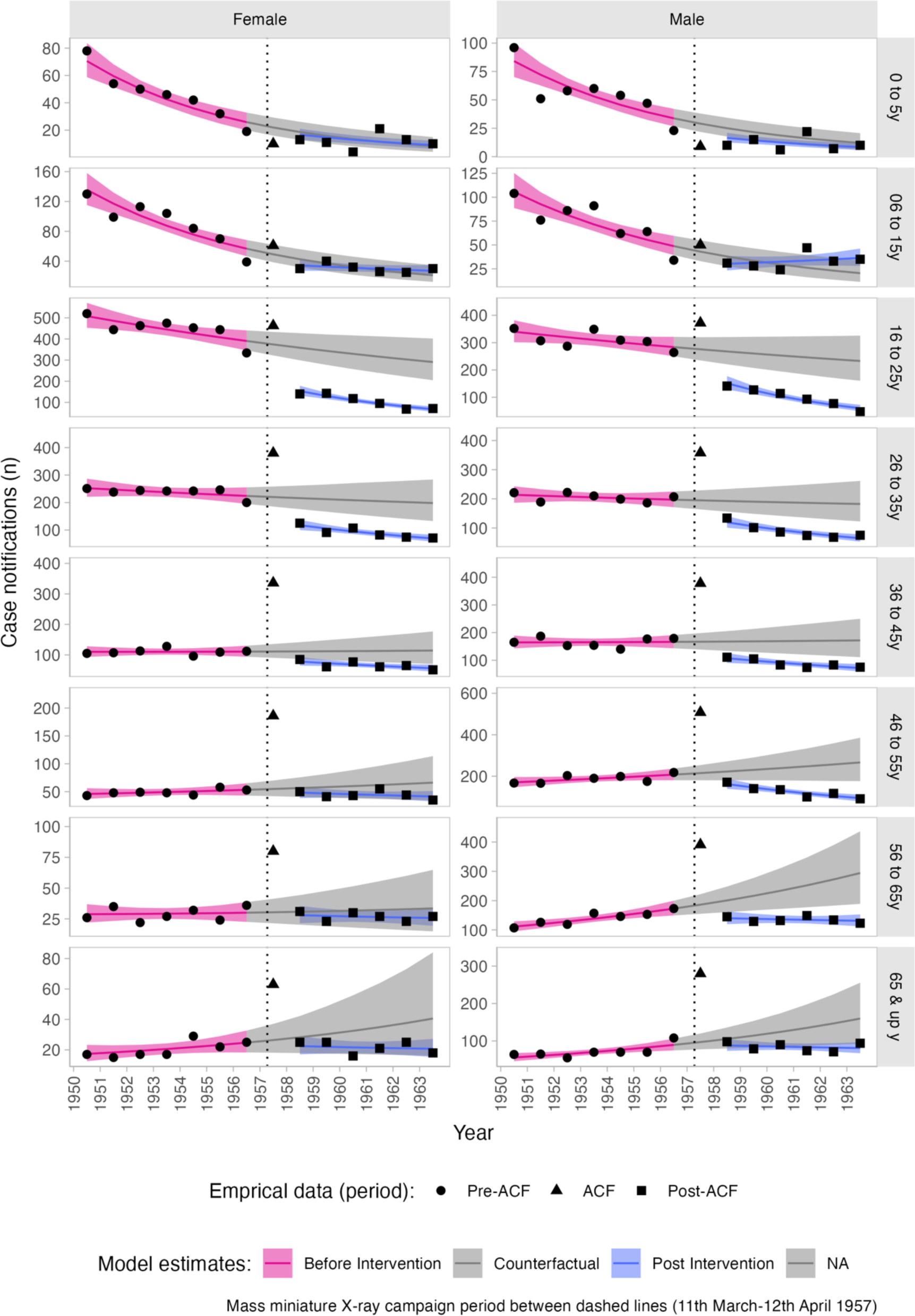
Impact of active case finding intervention on pulmonary tuberculosis by age and sex. Empirical and modelled case notification rates (per 100,000 population) by ward, with counterfactual of no active case finding intervention. The mass miniature X-ray active case finding campaign occurred between dashed lines (11th March-12th April 1957). CNR: case notification rate. ACF: active case finding.

